# Associations of vegetarianism with circulating lipids across varying genetic capacity: a cross-sectional Polygenic Score-by-Vegetarianism interaction study in UK Biobank

**DOI:** 10.64898/2026.04.29.26352068

**Authors:** Angela Ge, Yitang Sun, Alexandra Huong Chiang, Aryaman Singh, Huifang Xu, Kaixiong Ye

## Abstract

**Background:** Circulating lipid levels are influenced by both genetic and environmental factors. While vegetarianism has been linked to improved lipid profiles, it remains unclear whether these beneficial effects persist across individuals with varying genetic capacity for lipid metabolism.

**Objective:** We hypothesized that genetic capacity and vegetarianism interact to influence the circulating levels of four lipids, including total cholesterol (TC), low-density lipoprotein (LDL) cholesterol, high-density lipoprotein (HDL) cholesterol, and triglycerides (TG).

**Methods:** Our study included UK Biobank participants of European (EUR, n = 182,300), Central/South Asian (CSA, n = 2,627), African (n = 2,143), and East Asian (n = 1,031) ancestry. Utilizing polygenic scores (PGS) for four circulating lipids, we employed multivariable regression models to assess PGS-by-vegetarianism interactions for each lipid.

**Results:** Vegetarianism is associated with reduced levels of TC, LDL cholesterol, and HDL cholesterol, and with elevated levels of TG in the EUR cohort (p-value < 0.001). The same significant association patterns were observed for HDL cholesterol and TG in the CSA cohort. We did not detect significant PGS-by-vegetarianism interactions for any lipid traits (Interaction p-value > 0.05). There is a lack of evidence supporting that PGS modifies the associations between vegetarianism and lipid levels, nor that vegetarianism alters the effects of PGS on lipid levels.

**Conclusions:** Vegetarianism is associated with reduced TC, LDL cholesterol, and HDL cholesterol, as well as elevated TG among EUR participants, with similar patterns for HDL cholesterol and TG in CSA participants. These association effects of vegetarianism on circulating lipids are similar across individuals with varying genetic capacity for lipid metabolism.

## Introduction

In recent years, vegetarianism has gained popularity due to its associated health benefits as well as environmental, religious, cultural, and ethical motivations [1]. In particular, observational studies and randomized controlled trials indicate that vegetarian diets are associated with improved lipid profiles, reduced body mass index (BMI) and blood pressure, and enhanced insulin sensitivity [2–7], all of which are protective factors of cardiometabolic diseases [8]. Consistent with these improvements in risk profiles, vegetarianism has also been associated with reduced risk of type 2 diabetes, obesity, and cardiovascular disease (CVD) [2, 9–11]. With CVD being the leading cause of mortality in the United States, these associations of vegetarianism are particularly significant as dyslipidemia is a primary driver of CVD development [12].

Dyslipidemia is an abnormal lipid profile characterized by elevated circulating levels of total cholesterol (TC), low-density lipoprotein (LDL) cholesterol, and triglycerides (TG), as well as lowered circulating levels of high-density lipoprotein (HDL) cholesterol [13]. Blood lipid levels are influenced by environmental and lifestyle factors, including smoking, physical exercise, and dietary patterns (e.g., fish oil supplementation and vegetarian diets) [14–16]. Vegetarian diets are known to be associated with reduced levels of TC, LDL cholesterol, and HDL cholesterol [5, 17]; however, the findings for TG levels are inconclusive across studies. Some studies showed increased triglyceride levels for vegetarianism [17, 18], whereas others observed reductions [19, 20]. A meta-analysis concluded no association between vegetarianism and TG levels [6]. Therefore, further research is needed to resolve these inconsistencies.

Beyond environmental factors, genetic factors and their interaction with the environment can also influence circulating lipid levels. Circulating lipid levels are heritable, with estimated heritability ranging from 50% to 80% [21]. Previous genome-wide association studies (GWAS) have identified more than 175 genetic loci associated with the circulating levels of TC, LDL cholesterol, HDL cholesterol, and TG [22]. Some well-known loci include *APOB*, *APOE*, *LPL*, *PCSK9*, and *LIPC* [21, 23]. Notably, it has been shown that these genetic factors often modulate the relationship between circulating lipid levels and various lifestyle behaviors, such as physical activity [24], alcohol consumption [25], smoking [26], and fish oil supplementation [16, 27], through gene-by-environment interactions (G×E).

G×E occurs when genetic factors modify the effects of environmental exposures on the traits, or equivalently, when environmental exposures modify the effects of genetic factors on the traits [28]. For example, randomized controlled trials reported a significant interaction between *APOE* genotypes and diet on the plasma lipids [29, 30]. After 24 weeks on a low-glycemic index, lower-fat diet, *APOE* ε4 carriers showed a significantly greater reduction in TC when compared to *APOE* ε3 carriers [29]. Another G×E study found that SNP rs1042034 in *APOB* and SNP rs2072183 in *NPC1L1* modify the association between dietary cholesterol and plasma TC [31]. To date, few studies have investigated the gene-by-vegetarianism interaction on blood lipids. Our previous genome-wide interaction study analyzed 30 serum biomarkers and identified significant gene-by-vegetarianism interactions for calcium, testosterone, and estimated glomerular filtration rate [17]. However, no genome-wide significant gene-by-vegetarianism interactions were identified for lipid traits, possibly due to the limited power to detect small G×E effects at the level of individual genetic variants.

Polygenic score (PGS) aggregates the effects of individual trait-associated genetic variants into a single score to estimate one’s overall genetic risk for a disease or capacity for a trait [32]. PGS has shown potential to boost the power of variant discovery in GWAS [33] and improve disease prediction for clinical use [32]. By capturing the small effects of thousands of genetic loci simultaneously, PGS increases power and mitigates the multiple-testing burden inherent in single-variant association tests [34]. Therefore, a PGS-based framework may offer a more powerful approach for detecting interaction signals. In our recent G×E study using PGS as the genetic predictor, we found that fish oil supplementation significantly altered the associations between genetically predicted and observed lipid levels [16]. Other PGS-by-environment interaction (PGS×E) studies also demonstrated their feasibility, reporting significant interactions between PGS and lifestyle factors (e.g., BMI, diet, physical activity) for circulating cardiometabolic biomarkers and cardiovascular disease risk [35–39]. To our knowledge, no studies have examined PGS-by-vegetarianism interactions for blood lipids. It is important to evaluate whether vegetarianism interacts with PGS on blood lipids and to inform whether we should consider one’s genetic capacity when adopting vegetarianism to improve blood lipid profiles.

In this study, we hypothesize that there are interactions between PGS and vegetarianism on circulating blood lipid levels. We first examined the relationships between vegetarianism and circulating lipid levels among four ancestry groups, including European (EUR, n = 182,300), Central/South Asian (CSA, n = 2,627), African (AFR, n = 2,143), and East Asian (EAS, n = 1,031). PGS for TC, LDL cholesterol, HDL cholesterol, and TG in UK Biobank (UKB) participants were derived from a prior study [16]. We performed a PGS-by-vegetarianism interaction study to examine whether the overall genetic capacity modifies the associations between vegetarianism and circulating lipid levels. We also investigated whether these associations differ in participants with extreme PGS, focusing on participants in the highest and lowest PGS tertiles.

## Methods

### Study cohort

This study was conducted using the UKB, a prospective cohort comprising 502,369 participants, aged 37 to 73 years old at recruitment between 2006 and 2010 in the United Kingdom [40]. Ethical approval was obtained from the National Health Service North West - Haydock Research Ethics Committee, and all participants provided written consent before joining the study. The current study was approved under UKB project number 48818.

We conducted quality control for UKB participants. We excluded participants, whose self-reported sex and genetic sex did not match (n = 372), who withdrew consent as of 4/25/2023 (n = 10), who exhibited sex chromosome aneuploidy (n = 470), who were outliers in heterozygosity or with missing genotypes (n = 963), who demonstrated a high degree of genetic kinship to other participants (ten or more third-degree relatives) (n = 187). The genetic ancestry of participants was defined by the Pan-UK Biobank (Pan-UKB) Project, which assigned participants to one of six genetic ancestry groups, including EUR, CSA, AFR, EAS, Middle Eastern, and Admixed American [41]. We excluded participants of Middle Eastern and Admixed American (n = 2,605) ancestry due to their complex population structure and small sample sizes. We also excluded those who did not have genetic ancestry designations from Pan-UKB (n = 53,827). Additionally, our primary analysis was restricted to “verified” vegetarians based on two diet-related surveys and excluded participants who did not finish either survey (n = 255,555).

Blood samples were collected at recruitment for genotyping and biomarker measurements. Approximately 50,000 participants were genotyped using the BiLEVE array, and 450,000 participants were genotyped using the UKB Axiom array. Genetic imputation for all participants was performed using reference panels from the Haplotype Reference Consortium and UK10K [42].

### Assessment of blood lipid levels

Serum levels of TC, LDL cholesterol, HDL cholesterol, and TG were measured using standard hematological methodologies and quantified using Beckman Coulter AU5800 chemistry analyzer [43]. These blood lipid levels were standardized before regression analyses.

### Assessment of vegetarian status

We obtained participants’ vegetarian status from two surveys, including a touchscreen questionnaire from the initial visit and 24-hour dietary recalls at multiple follow-ups [17]. During the initial assessment center visit, most participants completed a questionnaire regarding their diet. Between 2009 and 2012, all participants were invited up to five times to fill out an online 24-hour dietary recall. A subset of participants, about 40% of the full cohort, filled out at least one dietary recall. In our designation of vegetarian, both vegans and vegetarians were included. We included participants who self-identified as vegetarian or vegan in the initial survey. To improve the accuracy of the vegetarian status, we defined “verified” vegetarians as participants who self-reported as vegetarian and also met the following criteria: 1) routinely follow a vegetarian or vegan diet (responded “Vegetarian diet (no meat, no poultry and no fish)” and/or “Vegan diet” to “Do you routinely follow a special diet?”), 2) have not had meat or poultry in the past 24 hours (responded “No” to “Did you eat any meat or poultry yesterday? Think about curry, stir-fry, sandwiches, pie fillings, sausages/burgers, liver, pate or mince”), 3) have not had fish or seafood in 24 hours (responded “No” to “Did you eat any fish or seafood yesterday? e.g. at breakfast, takeaway with chips, smoked fish, fish pate, tuna in sandwiches”), 4) have never eaten meat or fish (responded “Never” to all questions asking how often meat or fish was consumed), and 5) have not had any major diet changes in the past 5 years (responded “No” to “Have you made any major changes to your diet in the last 5 years?”).

### Source of polygenic scores

The PGS estimates an individual’s genetic capacity for a specific trait by aggregating the effects of genetic variants associated with the target trait [44]. In this study, PGS for four lipid traits (TC, LDL cholesterol, HDL cholesterol, and TG) were calculated by Sun *et al.* [16]. Full details on the calculation and validation of these scores have been previously described [16]. In brief, these PGS were derived from multi-ancestry GWAS meta-analyses conducted by Graham *et al.* within the Global Lipids Genetics Consortium. The primary GWAS studies included approximately 1.65 million individuals, with ancestry composition of 1,320,016 EUR, 146,492 EAS, 99,432 AFR, 40,963 CSA, and 48,057 Hispanic. In addition, the study incorporated GWAS from the UKB cohort, contributing 389,344 EUR-ancestry, 6,876 AFR-ancestry, and 6,827 CSA-ancestry participants. Our primary analysis focused on participants of EUR ancestry, utilizing PGS derived from GWAS that excluded UKB data to avoid bias from sample overlap. To assess the generalizability of our findings to non-EUR populations, we extended the analysis to participants of CSA and AFR ancestries. Furthermore, we conducted sensitivity analysis using alternative PGS based on GWAS from Graham *et al.* that included UKB (EUR, CSA, AFR, and EAS ancestries) and from Willer *et al.* (EUR only, n = 188,578) [45, 46]. For each lipid trait, PGS was standardized to one SD unit. Participants with missing PGS values (n = 279) were excluded from the analysis.

### Covariates

To account for possible confounding, we adjusted for sex, age, age^2^, genotyping array, assessment center, top 20 genetic principal components, BMI, Townsend deprivation index, statin use, smoking status, alcohol status, and physical activity. Sex and age at the time of recruitment were obtained through the central registry. The genotyping array indicated which array was used to genotype each participant. The assessment center was listed as the place where the participant consented to the study. The top 20 principal components were obtained from the Pan-UKB ancestry data. BMI was calculated by the participant’s height and weight upon the initial visit to the assessment center. The Townsend deprivation index, a measure of socioeconomic status, was evaluated for each participant at recruitment. Statin use was obtained through self-report during the initial assessment visit. Smoking and alcohol status were also gathered during the initial assessment, and participants were categorized as “never”, “previous”, or “current”. Physical activity was calculated using the International Physical Activity Questionnaire and participants were assigned to “low”, “moderate”, or “high” groups.

### Statistical analysis

We performed linear regression to examine the associations between vegetarianism and circulating lipid levels, adjusting for all covariates mentioned above. Individuals with missing observations in the covariates were removed. Analyses were conducted separately for verified and self-reported vegetarianism, and for different ancestries. We tested for G×E by incorporating a PGS-by-vegetarianism interaction term in our linear regression model. To interpret these interactions, we performed two types of stratified analyses. First, we compared participants in the highest and lowest PGS tertiles (i.e., top 33% and bottom 33%) to determine whether the impact of a vegetarian diet on blood lipid levels varies across genetic groups. Second, we stratified the participants into vegetarian and non-vegetarian groups to assess whether the association between PGS (i.e., genetic capacity) and the observed lipid phenotypes varies by dietary status.

All analyses were conducted using R (version 4.2.3).

## Results

### Participant characteristics

Our study cohort consisted of UKB participants of four ancestries: 182,300 EUR, 2,627 CSA, 2,143 AFR, and 1,031 EAS (**Figure 1**). Baseline participant characteristics are detailed in Table 1 and Supplementary Tables 1 and 2. Among the EUR participants, the mean age is 56 years, and 55% of participants are females. Among eligible participants of EUR, AFR, and EAS ancestries, approximately 1% were identified as verified vegetarians, while the proportion of self-reported vegetarians ranged from 3-4% (**Table 1**). Notably, CSA participants showed a larger proportion of verified vegetarians (9%) and self-reported vegetarians (25%). In both verified and self-reported EUR vegetarians, we observed a higher proportion of females, lower baseline levels of TC, LDL cholesterol, and TG, as well as higher baseline levels of HDL cholesterol, fewer smokers, and more individuals engaged in higher levels of physical activity (**Supplementary Tables 1 and 2**).

**Figure 1:**
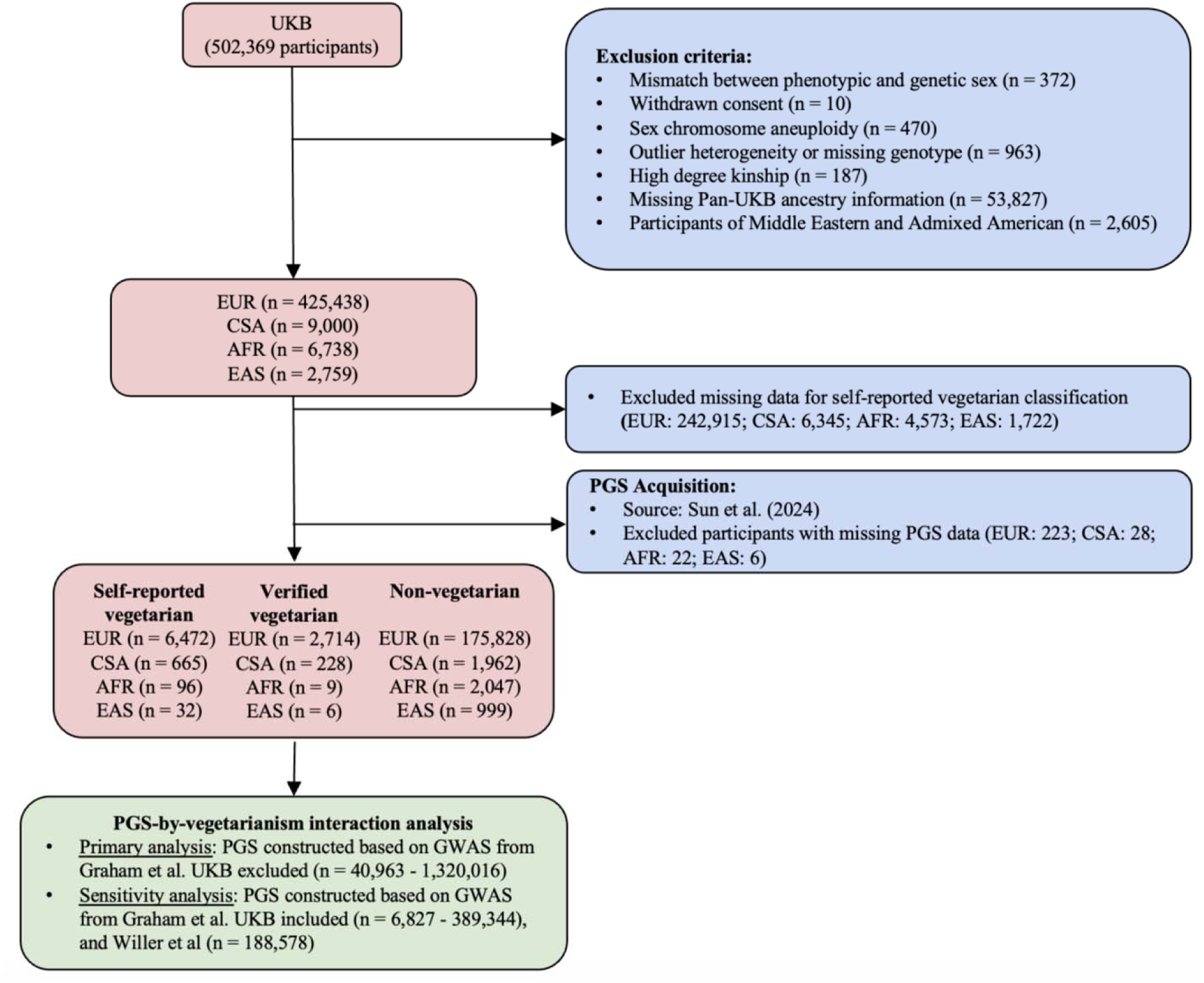
Summary of the study workflow. The participants of this study were from the UKB. Participant exclusions were applied, and PGS were derived for each participant based on existing GWAS. UKB, UK Biobank; Pan-UKB, Pan-ancestry genetic analysis of the UK Biobank; PGS, polygenic scores. EUR, European; CSA, Central/South Asian; AFR, African; EAS, East Asian.

**Table 1.** Characteristics of UK Biobank participants of European, Central/South Asian, African, and East Asian ancestries.

|  | **European**<br>**(n=182,300)** | **Central/South Asian**<br>**(n=2,627)** | **African**<br>**(n=2,143)** | **East Asian**<br>**(n=1,031)** |
| --- | --- | --- | --- | --- |
| **Age, years (SD)** | 56 (7.9) | 53 (8.4) | 51 (7.4) | 52 (7.7) |
| **Sex, female (%)** | 99,557 (55) | 1,210 (46) | 1,312 (61) | 711 (69) |
| **Body mass index, kg/m<sup>2</sup> (SD)** | 27 (4.6) | 27 (4.1) | 29 (5.4) | 24 (3.6) |
| **Total cholesterol, mmol/L (SD)** | 5.72 (1.12) | 5.36 (1.11) | 5.26 (1.06) | 5.64 (1.04) |
| **LDL cholesterol, mmol/L (SD)** | 3.57 (0.85) | 3.38 (0.84) | 3.27 (0.82) | 3.48 (0.78) |
| **HDL cholesterol, mmol/L (SD)** | 1.48 (0.39) | 1.29 (0.33) | 1.47 (0.37) | 1.5 (0.39) |
| **Triglycerides, mmol/L (SD)** | 1.69 (0.99) | 1.91 (1.13) | 1.13 (0.59) | 1.72 (1.03) |
| **Smoking status (%)** |  |  |  |  |
| Never | 102,581 (56) | 2,032 (77) | 1,520 (71) | 774 (75) |
| Previous | 65,474 (36) | 369 (14) | 402 (19) | 192 (19) |
| Current | 13,856 (8) | 209 (8) | 209 (10) | 61 (6) |
| **Alcohol status (%)** |  |  |  |  |
| Never | 4,620 (3) | 754 (29) | 272 (13) | 177 (17) |
| Previous | 5,412 (3) | 145 (6) | 102 (5) | 49 (5) |
| Current | 172,196 (94) | 1,712 (65) | 1,759 (82) | 805 (78) |
| **Physical activity (%)** |  |  |  |  |
| Low | 28,188 (15) | 512 (19) | 348 (16) | 180 (17) |
| Moderate | 65,518 (36) | 927 (35) | 656 (31) | 362 (35) |
| High | 60,861 (33) | 833 (32) | 716 (33) | 316 (31) |
| **Self-reported vegetarian, yes (%)** | 6,472 (4) | 665 (25) | 96 (4) | 32 (3) |
| **Verified vegetarian, yes (%)** | 2,714 (1) | 228 (9) | 9 (0) | 6 (1) |
| **Statin use, yes (%)** | 127,476 (70) | 1,739 (66) | 1,483 (69) | 548 (53) |

Abbreviation: SD: Standard deviation.

**Table 2.** Associations of PGS with the corresponding circulating lipids across vegetarian status in UKB participants of European and Central/South Asian ancestries.

| Lipids | Vegetarian |  |  | Non-vegetarian |  |  | P<sub>interaction</sub> |
| --- | --- | --- | --- | --- | --- | --- | --- |
|  | n | β (95% CI) | P-value | n | β (95% CI) | P-value |  |
| EUR |  |  |  |  |  |  |  |
| Total cholesterol | 2,275 | 0.296 (0.261, 0.331) | <2.0×10<sup>-16</sup> | 141,161 | 0.269 (0.264, 0.274) | <2.0×10<sup>-16</sup> | 0.265 |
| LDL cholesterol | 2,271 | 0.315 (0.280, 0.349) | <2.0×10<sup>-16</sup> | 140,899 | 0.288 (0.283, 0.293) | <2.0×10<sup>-16</sup> | 0.404 |
| HDL cholesterol | 2,071 | 0.236 (0.202, 0.270) | <2.0×10<sup>-16</sup> | 128,917 | 0.231 (0.226, 0.235) | <2.0×10<sup>-16</sup> | 0.720 |
| Triglycerides | 2,271 | 0.227 (0.188, 0.267) | <2.0×10<sup>-16</sup> | 141,076 | 0.260 (0.255, 0.264) | <2.0×10<sup>-16</sup> | 0.141 |
| CSA |  |  |  |  |  |  |  |
| Total cholesterol | 183 | 0.151 (0.010, 0.293) | 0.038 | 1,565 | 0.180 (0.134, 0.227) | 5.63×10<sup>-14</sup> | 0.464 |
| LDL cholesterol | 183 | 0.153 (0.000, 0.307) | 0.052 | 1,564 | 0.202 (0.156, 0.249) | <2.0×10<sup>-16</sup> | 0.183 |
| HDL cholesterol | 162 | 0.268 (0.146, 0.390) | 3.78×10<sup>-5</sup> | 1,434 | 0.190 (0.152, 0.227) | <2.0×10<sup>-16</sup> | 0.644 |
| Triglycerides | 183 | 0.279 (0.100, 0.459) | 0.003 | 1,565 | 0.252 (0.200, 0.305) | <2.0×10<sup>-16</sup> | 0.886 |

PGS was calculated using UKB-excluded GWAS summary statistics from Graham *et al.* Vegetarian status used a verified vegetarian classification. The number of vegetarians and non-vegetarians in the statistical analysis is represented by n. All lipid measurements were standardized to 1 SD unit. Abbreviation: 95% CI, 95% confident interval.

### Vegetarianism is significantly associated with all four blood lipids

We first tested the associations between vegetarianism and lipid levels for each of the four ancestry groups using multivariable regression models, adjusting for covariates including sex, age, age^2^, genotyping array, assessment center, top 20 genetic principal components, BMI, Townsend deprivation index, statin use, smoking status, alcohol status, and physical activity. Within the EUR sample, verified vegetarianism is significantly associated with decreased TC (β = -0.184, 95% CI = -0.224 – -0.144, p-value < 2.0×10^−16^), LDL cholesterol (β = -0.171, 95% CI = -0.211 – -0.130, p-value < 2.0×10^−16^), and HDL cholesterol levels (β = -0.165, 95% CI = - 0.201 – -0.129, p-value < 2.0×10^−16^), as well as significantly associated with increased triglyceride levels (β = 0.166, 95% CI = 0.127 – 0.204, p-value < 2.0×10^−16^) (**Figure 2, Supplementary Table 3**). We also found that verified vegetarianism is associated with reduced HDL cholesterol levels (β = -0.184, 95% CI = -0.315 – -0.054, p-value = 0.006) and increased TG levels (β = 0.244, 95% CI = 0.063 – 0.426, p-value = 0.008) in the CSA participants, which is consistent with the association patterns in the EUR group (**Figure 2**). Results from analyses using the self-reported vegetarian designation showed consistent patterns (**Supplementary Table 3**). Interestingly, self-reported vegetarianism in the CSA cohort is associated with decreased TC levels (β = -0.103, 95% CI = -0.205 – -0.001, p-value = 0.047). This finding may reflect the increased statistical power afforded by the larger sample size of self-reported vegetarians compared to that of verified vegetarians. In contrast, we did not observe any significant associations between vegetarianism and the levels of the four lipids in the AFR and EAS groups, likely due to the limited number of verified vegetarians.

**Figure 2:**
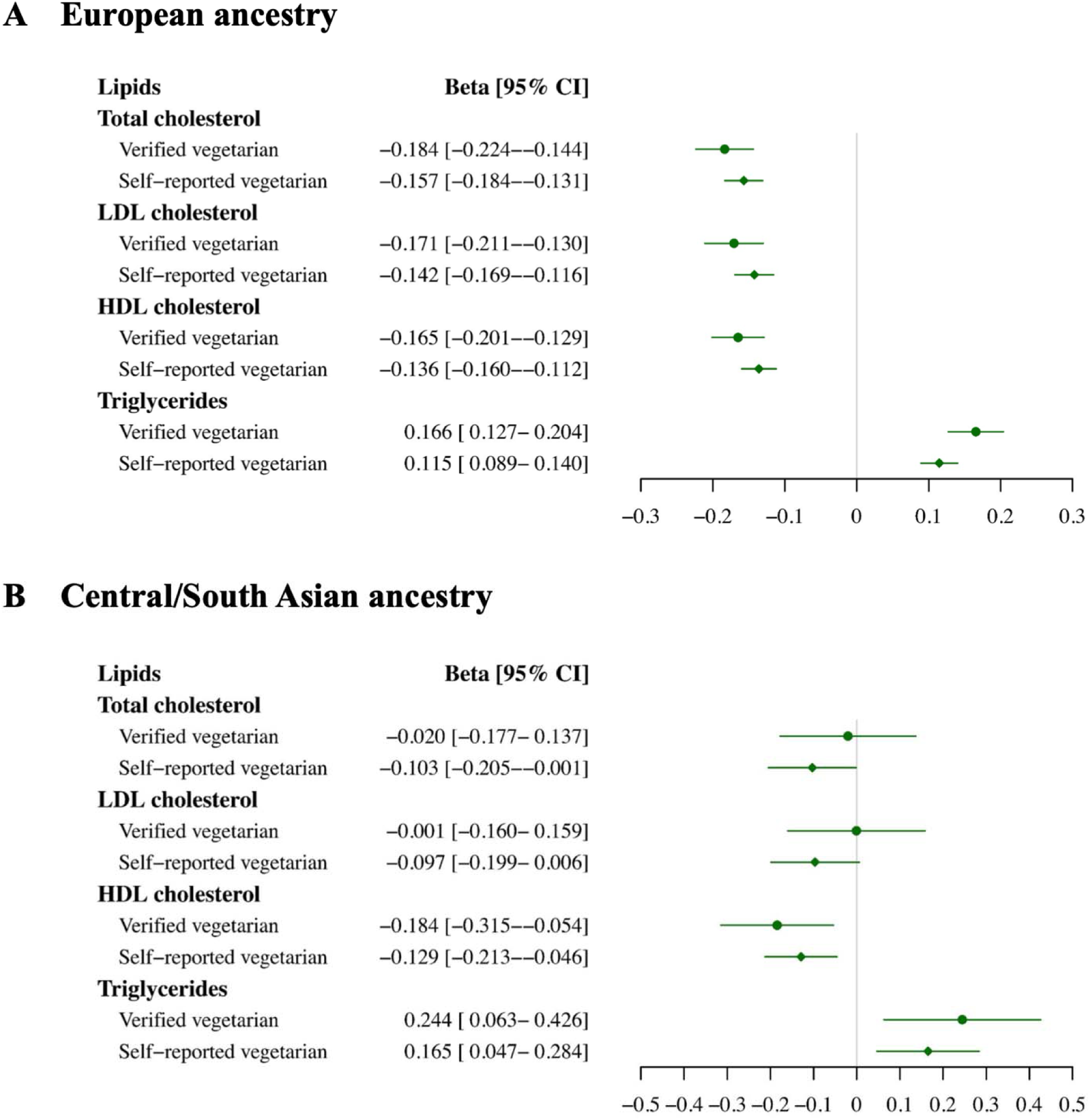
Associations of verified/self-reported vegetarians with observed four blood lipid levels. A: Result for participants of European ancestry. **B:** Result for participants of Central/South Asian ancestry. The beta value indicates the effect of verified/self-reported vegetarianism on the circulating level of each lipid. All lipid measurements were scaled to 1 SD unit. 95% CI, 95% confident interval.

### PGS are significantly predictive of the corresponding observed blood lipid levels

PGS for the four blood lipid levels were derived using GWAS summary statistics from Graham *et al*., which excluded UKB in the GWAS sample. The PGS were significantly associated with the corresponding lipid levels (p-value < 2.0×10^−16^) across the four ancestry groups (**Supplementary Table 4**). Among the EUR participants, a 1 SD increase in the respective PGS was associated with a 0.26 SD increase in TC levels (95% CI = 0.26 – 0.27), a 0.28 SD increase in LDL cholesterol (95% CI = 0.27 - 0.28), a 0.23 SD increase in HDL cholesterol (95% CI = 0.23 – 0.23), and a 0.26 SD increase in TG (95% CI = 0.26 – 0.26). PGS developed from both UKB-included GWAS from Graham *et al*. and another UKB-excluded GWAS from Willer *et al.* were also significantly predictive of their corresponding observed blood lipid levels (**Supplementary Table 4**).

### Absence of significant interactions between PGS and vegetarianism on blood lipid levels in the EUR population

To examine if there are interactions between lipid PGS and vegetarianism in influencing the observed lipid levels, we employed multivariable linear regression models incorporating a PGS-by-vegetarianism interaction term, PGS, vegetarianism, and all covariates mentioned above. The interaction analyses were performed only in the EUR and CSA cohorts, due to the limited numbers of vegetarians in the EAS and AFR ancestry groups.

We did not observe statistically significant interaction signals between PGS and vegetarianism for any of the four blood lipid levels. The associations between PGS and observed lipid levels were similar in vegetarians and non-vegetarians. For instance, among EUR participants, 1 SD increase in PGS was associated with a 0.296 SD increase in TC levels (95% CI = 0.261 - 0.331) in the verified vegetarian group, and a 0.269 SD increase (95% CI = 0.264 - 0.274) in the non-vegetarian group (interaction p-value = 0.265) (**Figure 3**, **Table 2**). Additionally, for vegetarians, per 1 SD increase, there was a 0.315 SD increase for LDL cholesterol (95% CI = 0.280 - 0.349), a 0.236 SD increase for HDL cholesterol (95% CI = 0.202- 0.270), and a 0.227 SD increase for TG (95% CI = 0.188 - 0.267). For non-vegetarians, the increase in lipid level per 1 SD increase in PGS was 0.288 SD (95% CI = 0.283 - 0.293), 0.231 SD (95% CI = 0.226 - 0.235), and 0.260 SD (95% CI = 0.255 - 0.264), for LDL cholesterol, HDL cholesterol, and TG levels, respectively. In summary, there is no evidence supporting that vegetarianism modifies the association between PGS and observed blood lipid levels.

**Figure 3:**
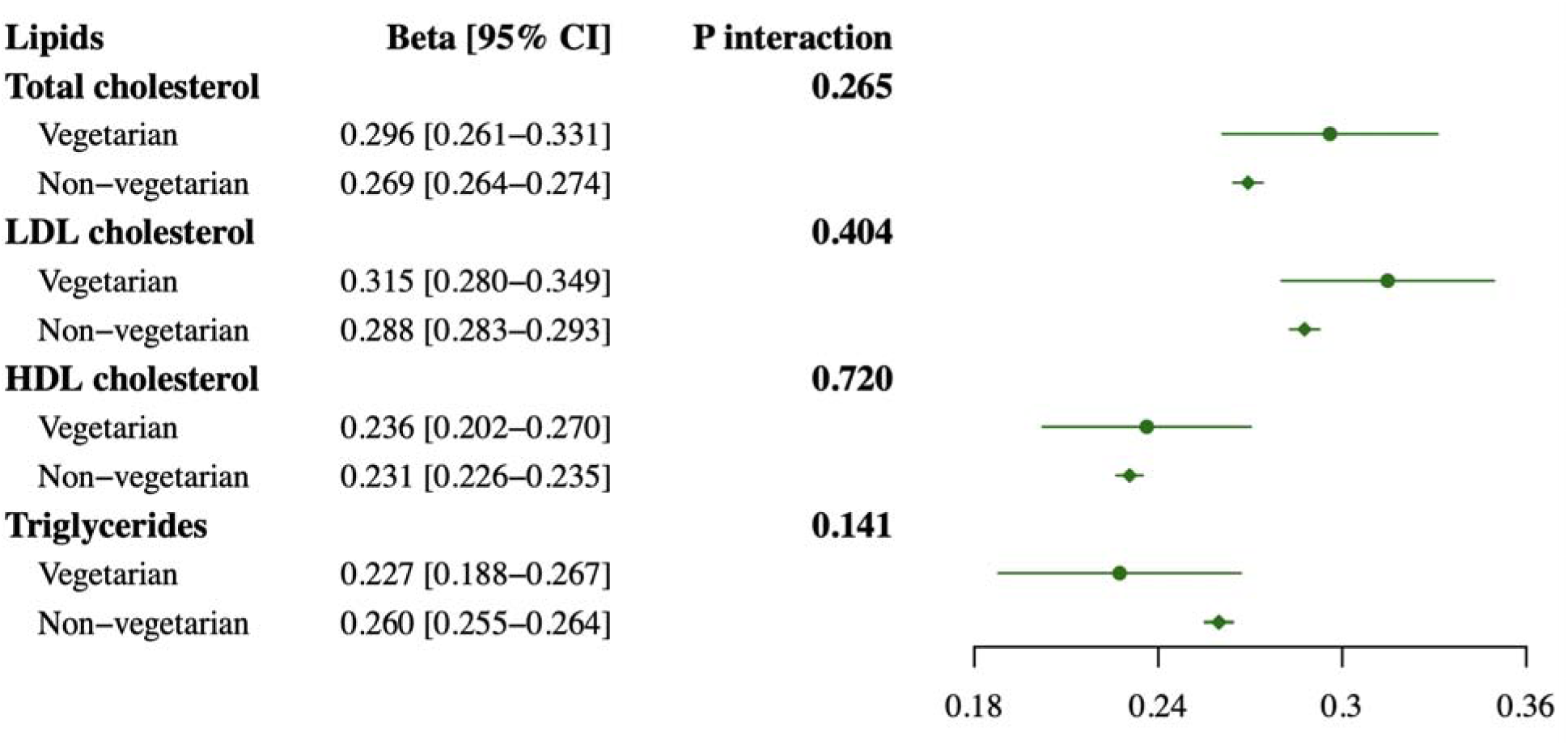
Associations of PGS with observed lipid levels stratified by verified vegetarian status for participants of European ancestry. This analysis was performed using PGS derived from the Graham *et al*. UKB-excluded GWAS. The beta value indicates the effect of PGS on the circulating level of each lipid in the vegetarian and non-vegetarian groups. All measurements were scaled to 1 SD unit. 95% CI, 95% confident interval.

We next focused on the two ends of the PGS distribution (top 33% vs bottom 33%) to investigate if the PGS can modify vegetarianism-lipid associations. Again, we applied multivariable linear regressions in a primary cohort of participants of EUR ancestry. No significant differences were observed in the association effects of vegetarianism on the four lipid levels between the high and low PGS groups (**Table 3**). For instance, the association effect of vegetarianism on TC is -0.146 (95% CI = -0.209 – -0.083) in the low PGS group and -0.177 (95% CI = -0.249 – -0.106) in the high PGS group (interaction p-value = 0.605). In summary, our analyses did not reveal evidence that genetic capacity modifies the effect of vegetarianism on circulating lipid levels.

**Table 3.** Association of vegetarianism (verified) with four blood lipid levels stratified by low and high PGS groups for the European and Central/South Asian cohorts.

| Lipids | Low PGS |  | High PGS |  | P<sub>interaction</sub> |
| --- | --- | --- | --- | --- | --- |
|  | β (95% CI) | P-value | β (95% CI) | P-value |  |
| EUR |  |  |  |  |  |
| Total cholesterol | -0.146 (-0.209, -0.083) | 5.66×10<sup>-6</sup> | -0.177 (-0.249, -0.106) | 1.32×10<sup>-6</sup> | 0.605 |
| LDL cholesterol | -0.169 (-0.233, -0.106) | 1.89×10<sup>-7</sup> | -0.198 (-0.270, -0.126) | 7.90×10<sup>-8</sup> | 0.384 |
| HDL cholesterol | -0.164 (-0.220, -0.108) | 1.05×10<sup>-8</sup> | -0.188 (-0.253, -0.123) | 1.83×10<sup>-8</sup> | 0.950 |
| Triglycerides | 0.162 (0.110, 0.213) | 6.42×10<sup>-10</sup> | 0.226 (0.147, 0.306) | 2.45×10<sup>-8</sup> | 0.491 |
| CSA |  |  |  |  |  |
| Total cholesterol | 0.059 (-0.208, 0.326) | 0.665 | -0.090 (-0.384, 0.204) | 0.55 | 0.284 |
| LDL cholesterol | 0.117 (-0.157, 0.392) | 0.403 | 0.045 (-0.258, 0.349) | 0.771 | 0.352 |
| HDL cholesterol | -0.139 (-0.334, 0.055) | 0.161 | -0.186 (-0.428, 0.057) | 0.134 | 0.634 |
| Triglycerides | 0.297 (0.035, 0.559) | 0.027 | 0.178 (-0.240, 0.597) | 0.404 | 0.682 |

PGS was calculated using GWAS summary statistics from Graham *et al.* excluding UKB. Vegetarian status used a verified vegetarian classification. The beta value indicates the effect of vegetarianism on each blood lipid level in low (bottom 33%) and high (top 33%) PGS groups. Vegetarian status used a verified vegetarian classification. Abbreviations: EUR, European; CSA, Central/South Asian; 95% CI, 95% confident interval.

### Absence of interaction signals in the CSA population

Due to the small sample sizes of vegetarians in the EAS and AFR groups, we restricted additional interaction analysis to the CSA group. No significant interaction was detected between PGS and verified vegetarianism for any of the four blood lipid levels in the CSA group (interaction p-value > 0.05), consistent with the results in the EUR group (**Table 2**). Additional analyses restricting to the top 33% and bottom 33% of the PGS distribution, similarly, did not find evidence for interaction (**Table 3**).

### Sensitivity analyses with self-reported vegetarianism and alternative PGS

To ensure the robustness of our findings, we conducted sensitivity analyses that 1) utilized alternative PGS derived from UKB-included GWAS from Graham *et al*. and another GWAS from Willer *et al.*, and 2) repeated interaction analyses using self-reported vegetarian definitions (**Supplemental Figures 1-2, Supplementary Tables 4-8**). Overall, these sensitivity analyses revealed consistent results for the circulating levels of TC, LDL cholesterol, and HDL cholesterol in terms of the significant associations of PGS and vegetarianism, independently, with the observed lipid levels and the lack of interaction signals between PGS and vegetarianism. Interestingly, we detected significant interaction between PGS, developed from the Willer *et al.* study, and vegetarianism in influencing TG levels, for both verified (interaction p-value = 0.015) and self-reported vegetarianism (interaction p-value = 0.008) (**Supplementary Figures 1-2 and Supplementary Tables 5-6**). Verified vegetarianism attenuates the association effect of PGS on TG (β = 0.155, 95% CI: 0.116 – 0.195, p-value = 1.71×10^−14^) compared to non-vegetarian (β = 0.206, 95% CI: 0.201 – 0.210, p-value = 2.0×10^−13^; **Supplementary Tables 5**). Additionally, in participants of EUR ancestry, vegetarianism (self-reported) has stronger association effect on TG level (interaction p-value = 0.004) in the low PGS group (β = 0.139, 95% CI: 0.103 – 0.176, p-value = 5.52×10^−14^) than in the high PGS group (β = 0.116, 95% CI: 0.066 – 0.166, p-value = 5.39×10^−6^) (**Supplementary Table 8**). However, this significant PGS-by-vegetarianism interaction on TG was not robust to the usage of other PGS derivation approaches.

## Discussion

Our study investigated the associations of vegetarianism and PGS with four blood lipid levels and tested whether vegetarianism and PGS interact to influence these levels. We found that vegetarianism was significantly associated with lower levels of TC, LDL cholesterol, and HDL cholesterol, along with increased TG levels in the EUR population. Consistent association patterns were also observed in the participants of CSA ancestry. We did not consistently detect significant PGS-by-vegetarianism interaction signals. In other words, we did not have sufficient evidence to support that genetic capacity for blood lipid levels modifies the associations between vegetarianism and blood lipid levels, or that the vegetarian status modifies the associations between genetically predicted (i.e., PGS) and observed blood lipid levels.

Our study revealed that vegetarianism is negatively associated with TC, LDL cholesterol, and HDL cholesterol levels, while positively associated with TG levels. These patterns are consistent with some previous reports [5, 17]. Existing literature regarding the effects of vegetarianism on TG has been inconsistent. One cross-sectional study on 76 participants showed that vegetarianism significantly lowered TG levels, while another cross-sectional study on 187 children showed that vegetarianism had TG-increasing effects [19, 47]. In contrast, a meta-analysis of randomized controlled trials observed insignificant results for vegetarianism’s effects on TG [6]. However, our study included a larger sample size (2,714 vegetarians and 175,828 non-vegetarians), controlled for potential confounders, provided greater statistical power, and supported evidence for the positive relationship between vegetarianism and TG levels. We also observed a consistent association pattern for HDL cholesterol and TG levels in the CSA cohort. Therefore, our study added to the evidence that vegetarianism is associated with a decrease in TC, HDL cholesterol, and LDL cholesterol, as well as an increase in TG.

There is extensive literature regarding the potential biological mechanisms behind vegetarianism’s effects on lipid levels. Significantly decreased TC, LDL cholesterol, and HDL cholesterol levels in vegetarians are likely due to reduced intake of saturated fats in meat and animal products, while dietary saturated fats are an established factor in raising circulating LDL cholesterol levels [48]. In addition, replacing dietary saturated fats with polyunsaturated fats effectively lowers LDL cholesterol level and incidence of CVD [48]. Decreased cholesterol levels in vegetarians have also been ascribed to higher intake of dietary fiber that can reduce cholesterol reabsorption and inflammation, as well as cholesterol-lowering compounds such as plant sterols/stanols [49, 50]. Despite the conflicting results on the relationship between TG and vegetarianism in existing meta-analyses, we observed increased TG levels in vegetarians, which may be due to increased simple carbohydrate intake and decreased vitamin D intake [17, 51].

Our study did not find any interactions between PGS and vegetarianism on influencing blood lipid levels in our primary analysis, which might be explained by the following reasons. First, there is indeed no significant interaction between PGS and vegetarianism on blood lipids. While both PGS and vegetarianism are associated with blood lipids, their effects are independent of each other. Second, given the fact that the sample size required to detect G×E is much larger than that of GWAS signals, the sample size of vegetarians in this study (i.e., n = 2,714 in EUR, n = 228 in CSA) may not be sufficient to detect modest interaction effects [52]. Additionally, the coarsened, binarized classification of vegetarianism also limits the statistical power [34]. Third, the PGS is constructed by summing up genetic variants weighted by marginal main effects without considering interaction effects. It is possible that some variants that interact with vegetarianism fail to be identified, thereby leading to a null PGS-by-vegetarianism interaction [53]. Evidence already shows that there are interactions between individual variants (e.g., rs1042034 in *APOB*, rs2072183 in *NPC1L1*, and rs380867 in *CYP7A1*) and plant-based diets on blood lipid levels [29, 30]. Lastly, the fundamental P+T method adopted in this study is a basic PGS approach. Other PGS methods, such as those considering functional annotations, incorporating multi-ancestry, and especially those considering interaction effects, may improve performance and produce different results [53, 54]. Surprisingly, our sensitivity analysis found a significant interaction between PGS derived from the Willer *et al*. study and vegetarianism on TG levels. The GWAS from Willer *et al*. was an earlier study with a smaller sample size and was included in the Graham *et al*. meta-analysis. Given the smaller discovery sample size and the consistent absence of interaction signals across four ancestral groups, we suspect this might be a random false positive. Further inclusion of larger samples, different PGS approaches, and more precise, quantitative data on the types and duration of vegetarian diets is needed to verify the presence or absence of PGS-by-vegetarianism interactions on blood lipid levels.

Our study has multiple strengths. First, our sample size for the EUR cohort is very large compared to existing studies. Second, we used the large and most recent GWAS summary statistics from various studies for our primary and sensitivity analyses to ensure the robustness of our findings. Third, we extended our study to non-European populations and found consistent results in the CSA cohort. Additionally, while existing studies have analyzed genetic and environmental effects on lipid levels distinctly, our study is the first of its kind to investigate the PGS-by-vegetarianism interaction (i.e., considering an individual’s overall genetic capacity) on blood lipid traits.

Several limitations should be considered. First, our interaction analysis had a primary focus on participants of EUR ancestry, which may make it difficult to generalize results to other ancestry groups. This was due to the limited sample sizes of the other ancestry groups in the interaction analysis. Second, a common limitation for studies of vegetarianism is the arbitrary nature of the term “vegetarian” which may result in possible discrepancies in vegetarian status. However, we did utilize a rigorous categorization method to determine verified vegetarian status based on two diet-related surveys. Sensitivity analyses utilizing the self-reported vegetarian status and larger sample sizes revealed consistent results. Third, our findings may be subject to unaccounted confounding, including covariates that possibly influence individuals’ choice to become vegetarian, as well as other dietary habits often associated with vegetarianism. Lastly, we cannot fully rule out reverse causation in our study. For example, some participants may adopt a vegetarian lifestyle in response to observed unhealthy lipid levels.

## Conclusion

Our study shows that vegetarianism is associated with reduced circulating levels of TC, LDL cholesterol, and HDL cholesterol, along with elevated TG levels in participants of EUR ancestry. The same association patterns were observed for the levels of HDL cholesterol and TG among CSA participants. We do not have sufficient evidence to support the interaction between vegetarianism and the genetic capacity of circulating lipids. Our findings suggest that vegetarianism has the same effects on blood lipid levels across individuals with varying genetic capacity for lipid metabolism.

## Abbreviations

AFR: African
BMI: body mass index
CSA: Central/South Asian
CVD: cardiovascular disease
EAS: East Asian
EUR: European
GWAS: genome-wide association studies
G×E: gene-by-environment interaction
HDL: high-density lipoprotein
LDL: low-density lipoprotein
Pan-UKB: Pan-UK Biobank
PGS: polygenic score
SNP: single nucleotide polymorphism
UKB: UK Biobank;
SD: Standard deviation.

## Acknowledgements

This research has been conducted using the UK Biobank resource under Application Number 48818. We would like to thank all participants for their contribution to science. We thank the Global Lipids Genetics Consortium for providing access to genome-wide association study summary statistics. We also want to thank the University of Georgia GACRC staff for facilitating our data analysis.

YS, HX, and KY designed the research; AG, YS, HX, and KY conducted the research; AG, YS, AHC, and AS analyzed data; AG and YS wrote the first draft of the manuscript; HX and KY critically revised the manuscript; All authors read and approved the final manuscript; All authors had primary responsibility for the final content.

## Data Availability

GWAS summary statistics from Graham *et al*. were downloaded from https://csg.sph.umich.edu/willer/public/glgc-lipids2021/. GWAS summary statistics from Willer *et al*. were downloaded from https://csg.sph.umich.edu/willer/public/lipids2013/. All of the code for this study are publicly accessible via https://github.com/angelage678/GxE_vegetarianism_lipids.

## Supplementary Figures

**Supplementary Figure 1:**
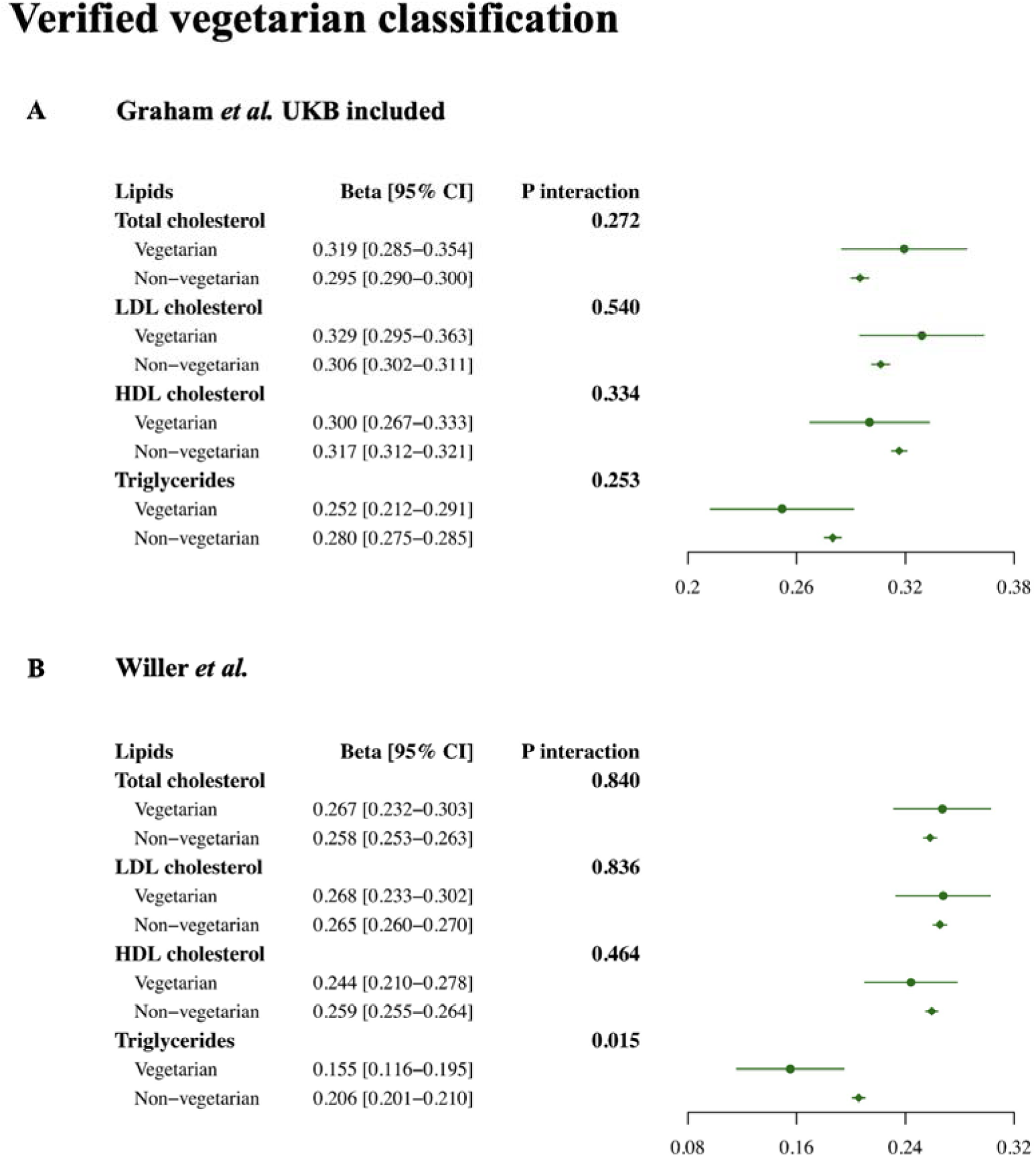
Association of PGS with observed lipid levels stratified by verified vegetarian status for participants of European ancestry (sensitivity analyses). Analyses were performed using **A)** PGS based on the UKB-included GWAS from Graham *et al*., and **B)** PGS based on the GWAS from Willer *et al*. dataset. The beta value indicates the effect of PGS on each blood lipid level in verified vegetarian and non-vegetarian groups, respectively. All lipid measurements were scaled to 1 SD unit. Abbreviation: 95% CI, 95% confident interval.

**Supplementary Figure 2.**
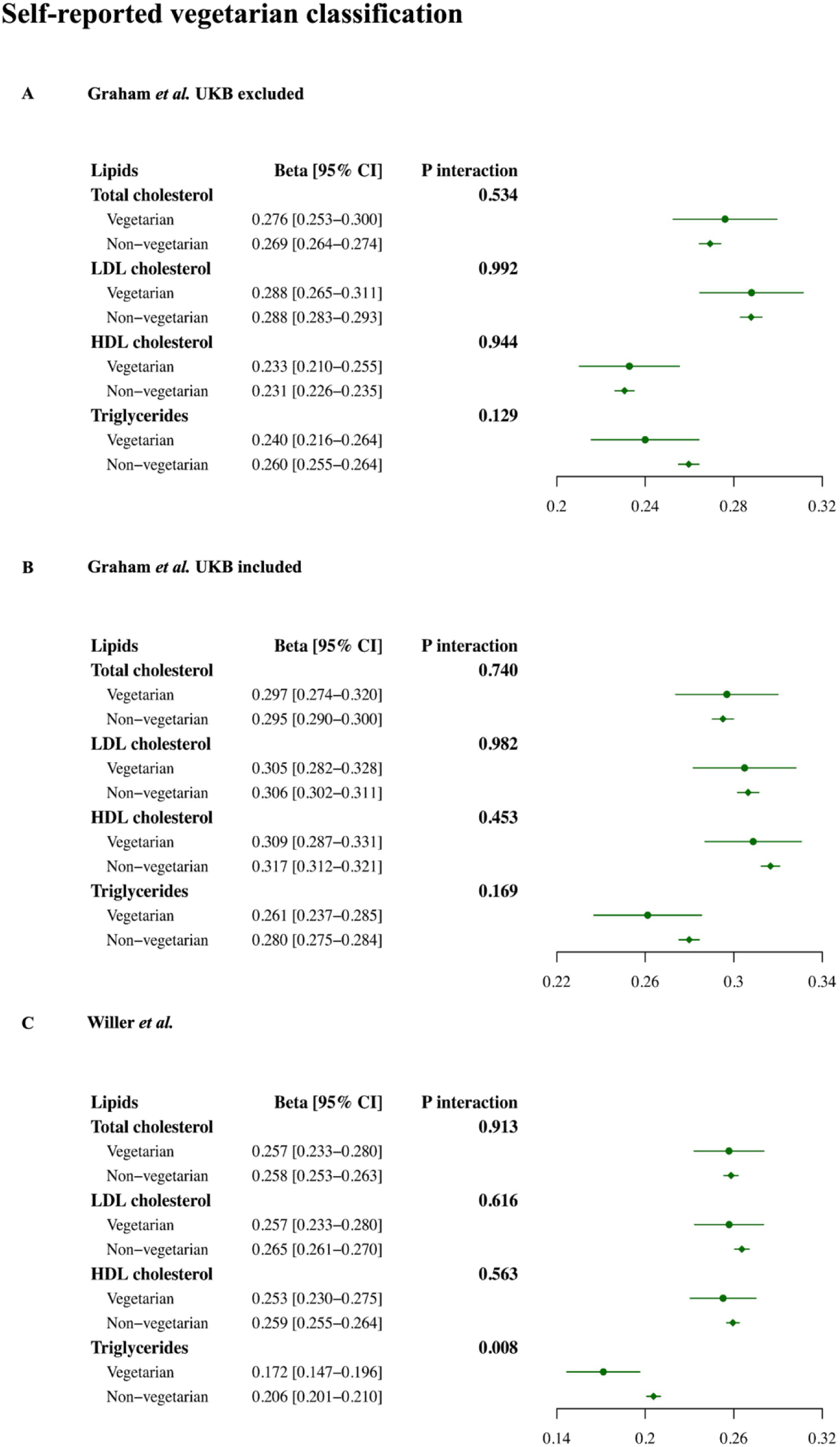
Association of PGS with observed lipid levels stratified by self-reported vegetarian status for participants of European ancestry (sensitivity analyses). Analyses were performed using **A:** PGS based on the GWAS from Graham *et al*. that excludes the UKB dataset, **B)** PGS based on the GWAS from Graham *et al*. that includes the UKB dataset, and **C)** PGS based on the GWAS from Willer *et al*. dataset. The beta value indicates the effect of PGS on each blood lipid level in self-reported vegetarian and non-vegetarian groups, respectively. All lipid measurements were scaled to 1 SD unit. Abbreviation: 95% CI, 95% confident interval.

## Supplementary Tables

**Supplementary Table 1.**
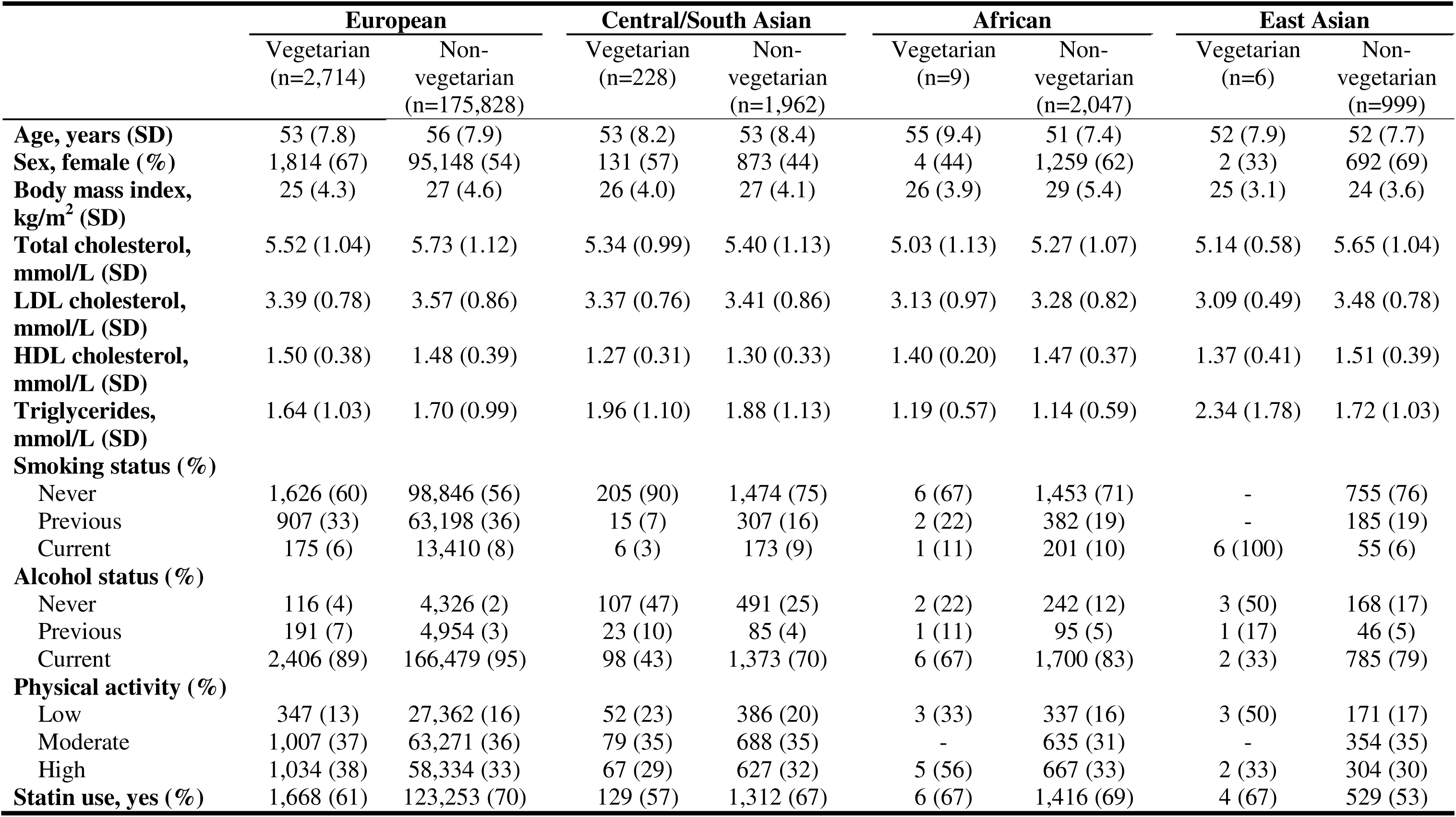
Characteristics of UK Biobank participants of European, Central/South Asian, African, and East Asian ancestries stratified by verified vegetarian status. Values are numbers (%) for categorical variables and mean (SD) for continuous variables. Abbreviation: SD, Standard deviation.

**Supplementary Table 2.**
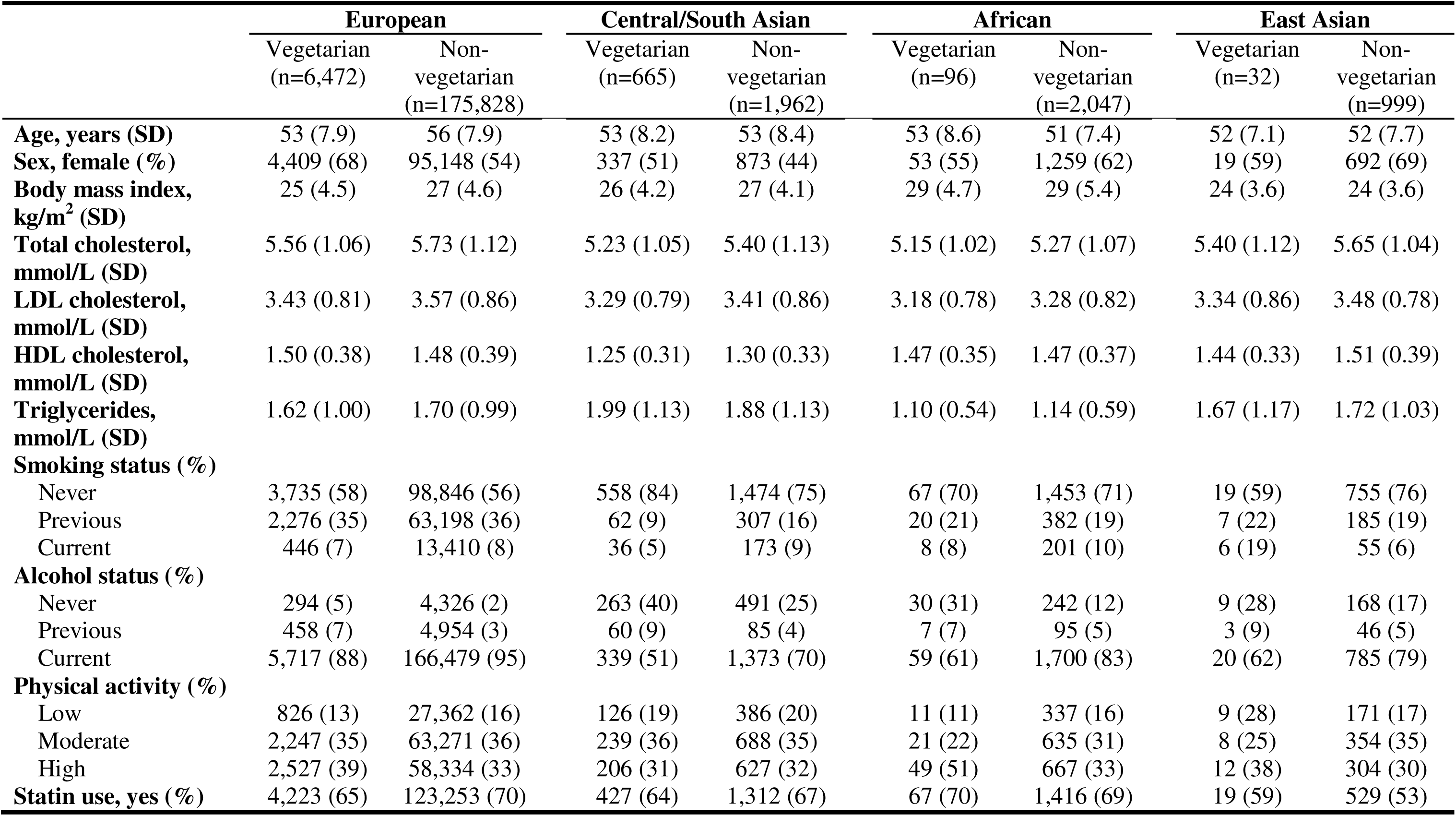
Characteristics of UK Biobank participants for European, Central/South Asian, African, and East Asian ancestries stratified by self-reported vegetarian status. Values are numbers (%) for categorical variables and mean (SD) for continuous variables. Abbreviation: SD, Standard deviation.

|  | **European** |  | **Central/South Asian** |  | **African** |  | **East Asian** |  |
| --- | --- | --- | --- | --- | --- | --- | --- | --- |
|  | Vegetarian<br>(n=6,472) | Non-vegetarian<br>(n=175,828) | Vegetarian<br>(n=665) | Non-vegetarian<br>(n=1,962) | Vegetarian<br>(n=96) | Non-vegetarian<br>(n=2,047) | Vegetarian<br>(n=32) | Non-vegetarian<br>(n=999) |
| **Age, years (SD)** | 53 (7.9) | 56 (7.9) | 53 (8.2) | 53 (8.4) | 53 (8.6) | 51 (7.4) | 52 (7.1) | 52 (7.7) |
| **Sex, female (%)** | 4,409 (68) | 95,148 (54) | 337 (51) | 873 (44) | 53 (55) | 1,259 (62) | 19 (59) | 692 (69) |
| **Body mass index, kg/m<sup>2</sup> (SD)** | 25 (4.5) | 27 (4.6) | 26 (4.2) | 27 (4.1) | 29 (4.7) | 29 (5.4) | 24 (3.6) | 24 (3.6) |
| **Total cholesterol, mmol/L (SD)** | 5.56 (1.06) | 5.73 (1.12) | 5.23 (1.05) | 5.40 (1.13) | 5.15 (1.02) | 5.27 (1.07) | 5.40 (1.12) | 5.65 (1.04) |
| **LDL cholesterol, mmol/L (SD)** | 3.43 (0.81) | 3.57 (0.86) | 3.29 (0.79) | 3.41 (0.86) | 3.18 (0.78) | 3.28 (0.82) | 3.34 (0.86) | 3.48 (0.78) |
| **HDL cholesterol, mmol/L (SD)** | 1.50 (0.38) | 1.48 (0.39) | 1.25 (0.31) | 1.30 (0.33) | 1.47 (0.35) | 1.47 (0.37) | 1.44 (0.33) | 1.51 (0.39) |
| **Triglycerides, mmol/L (SD)** | 1.62 (1.00) | 1.70 (0.99) | 1.99 (1.13) | 1.88 (1.13) | 1.10 (0.54) | 1.14 (0.59) | 1.67 (1.17) | 1.72 (1.03) |
| **Smoking status (%)** |  |  |  |  |  |  |  |  |
| Never | 3,735 (58) | 98,846 (56) | 558 (84) | 1,474 (75) | 67 (70) | 1,453 (71) | 19 (59) | 755 (76) |
| Previous | 2,276 (35) | 63,198 (36) | 62 (9) | 307 (16) | 20 (21) | 382 (19) | 7 (22) | 185 (19) |
| Current | 446 (7) | 13,410 (8) | 36 (5) | 173 (9) | 8 (8) | 201 (10) | 6 (19) | 55 (6) |
| **Alcohol status (%)** |  |  |  |  |  |  |  |  |
| Never | 294 (5) | 4,326 (2) | 263 (40) | 491 (25) | 30 (31) | 242 (12) | 9 (28) | 168 (17) |
| Previous | 458 (7) | 4,954 (3) | 60 (9) | 85 (4) | 7 (7) | 95 (5) | 3 (9) | 46 (5) |
| Current | 5,717 (88) | 166,479 (95) | 339 (51) | 1,373 (70) | 59 (61) | 1,700 (83) | 20 (62) | 785 (79) |
| **Physical activity (%)** |  |  |  |  |  |  |  |  |
| Low | 826 (13) | 27,362 (16) | 126 (19) | 386 (20) | 11 (11) | 337 (16) | 9 (28) | 171 (17) |
| Moderate | 2,247 (35) | 63,271 (36) | 239 (36) | 688 (35) | 21 (22) | 635 (31) | 8 (25) | 354 (35) |
| High | 2,527 (39) | 58,334 (33) | 206 (31) | 627 (32) | 49 (51) | 667 (33) | 12 (38) | 304 (30) |
| **Statin use, yes (%)** | 4,223 (65) | 123,253 (70) | 427 (64) | 1,312 (67) | 67 (70) | 1,416 (69) | 19 (59) | 529 (53) |

**Supplementary Table 3.**
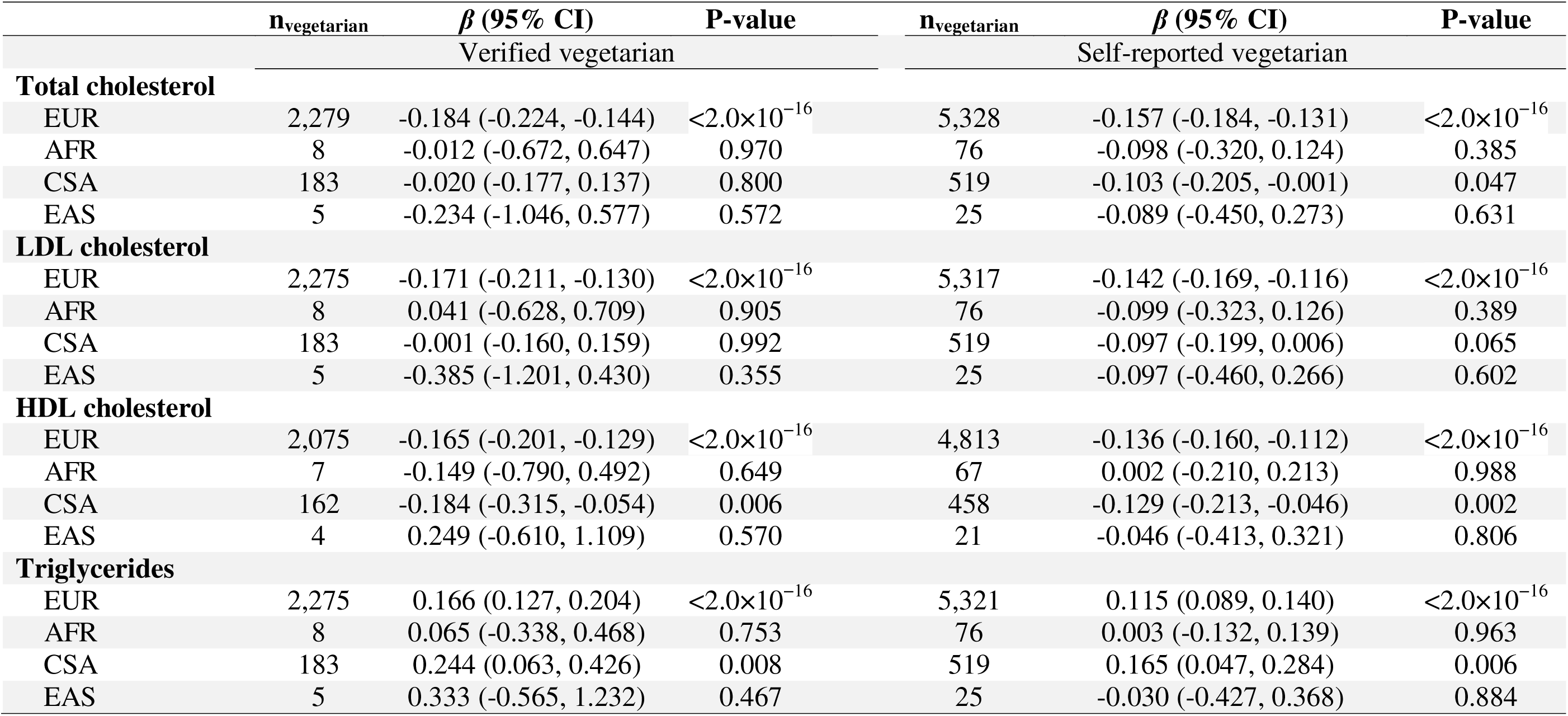
Effect of verified/self-reported vegetarianism on four blood lipid levels in participants of European, African, Central/South Asian, and East Asian ancestries. The number of verified/self-reported vegetarians was represented by n_vegetarian_. Abbreviation: 95% CI, 95% confident interval; EUR, European; CSA, Central/South Asian; AFR, African; EAS, East Asian.

**Supplementary Table 4.**
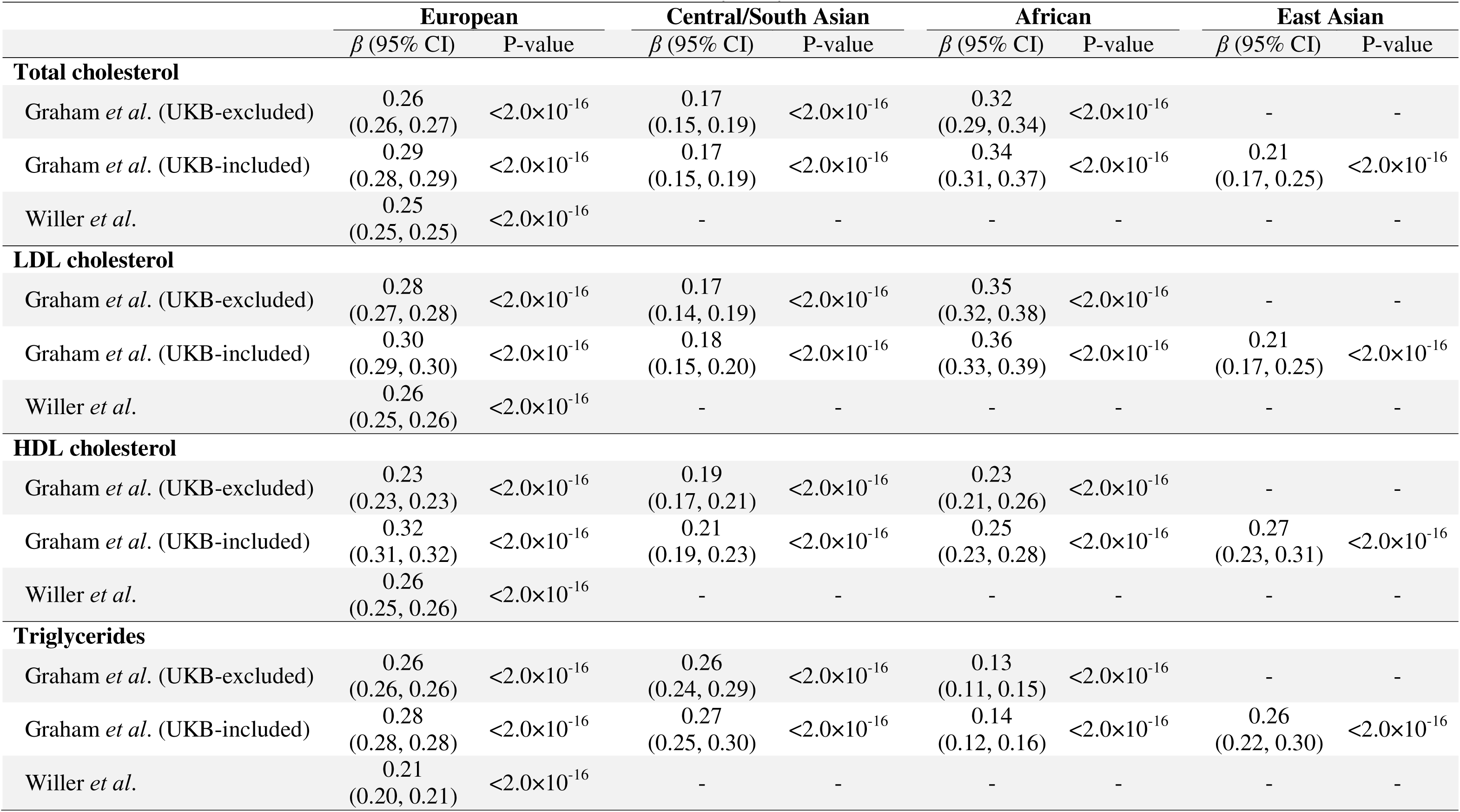
Associations of lipid PGS with the corresponding circulating lipid in UKB participants of European, Central/South Asian, African, and East Asian ancestries (sensitivity analyses). PGS were calculated using UKB-excluded and UKB-included GWAS from Graham *et al*., and GWAS from Willer *et al*, respectively. The beta value indicates the effect of PGS on each blood lipid level. Some results are indicated as “-” due to unavailable data in that analysis. Abbreviations: EUR, European; CSA, Central/South Asian; AFR, African; EAS, East Asian; 95% CI, 95% confident interval; PGS, polygenic scores

|  | **European** |  | **Central/South Asian** |  | **African** |  | **East Asian** |  |
| --- | --- | --- | --- | --- | --- | --- | --- | --- |
| | $\beta$ (95% CI) | P-value | $\beta$ (95% CI) | P-value | $\beta$ (95% CI) | P-value | $\beta$ (95% CI) | P-value |
| **Total cholesterol** |  |  |  |  |  |  |  |  |
| Graham *et al.* (UKB-excluded) | 0.26<br>(0.26, 0.27) | <2.0×10<sup>-16</sup> | 0.17<br>(0.15, 0.19) | <2.0×10<sup>-16</sup> | 0.32<br>(0.29, 0.34) | <2.0×10<sup>-16</sup> | - | - |
| Graham *et al.* (UKB-included) | 0.29<br>(0.28, 0.29) | <2.0×10<sup>-16</sup> | 0.17<br>(0.15, 0.19) | <2.0×10<sup>-16</sup> | 0.34<br>(0.31, 0.37) | <2.0×10<sup>-16</sup> | 0.21<br>(0.17, 0.25) | <2.0×10<sup>-16</sup> |
| Willer *et al.* | 0.25<br>(0.25, 0.25) | <2.0×10<sup>-16</sup> | - | - | - | - | - | - |
| **LDL cholesterol** |  |  |  |  |  |  |  |  |
| Graham *et al.* (UKB-excluded) | 0.28<br>(0.27, 0.28) | <2.0×10<sup>-16</sup> | 0.17<br>(0.14, 0.19) | <2.0×10<sup>-16</sup> | 0.35<br>(0.32, 0.38) | <2.0×10<sup>-16</sup> | - | - |
| Graham *et al.* (UKB-included) | 0.30<br>(0.29, 0.30) | <2.0×10<sup>-16</sup> | 0.18<br>(0.15, 0.20) | <2.0×10<sup>-16</sup> | 0.36<br>(0.33, 0.39) | <2.0×10<sup>-16</sup> | 0.21<br>(0.17, 0.25) | <2.0×10<sup>-16</sup> |
| Willer *et al.* | 0.26<br>(0.25, 0.26) | <2.0×10<sup>-16</sup> | - | - | - | - | - | - |
| **HDL cholesterol** |  |  |  |  |  |  |  |  |
| Graham *et al.* (UKB-excluded) | 0.23<br>(0.23, 0.23) | <2.0×10<sup>-16</sup> | 0.19<br>(0.17, 0.21) | <2.0×10<sup>-16</sup> | 0.23<br>(0.21, 0.26) | <2.0×10<sup>-16</sup> | - | - |
| Graham *et al.* (UKB-included) | 0.32<br>(0.31, 0.32) | <2.0×10<sup>-16</sup> | 0.21<br>(0.19, 0.23) | <2.0×10<sup>-16</sup> | 0.25<br>(0.23, 0.28) | <2.0×10<sup>-16</sup> | 0.27<br>(0.23, 0.31) | <2.0×10<sup>-16</sup> |
| Willer *et al.* | 0.26<br>(0.25, 0.26) | <2.0×10<sup>-16</sup> | - | - | - | - | - | - |
| **Triglycerides** |  |  |  |  |  |  |  |  |
| Graham *et al.* (UKB-excluded) | 0.26<br>(0.26, 0.26) | <2.0×10<sup>-16</sup> | 0.26<br>(0.24, 0.29) | <2.0×10<sup>-16</sup> | 0.13<br>(0.11, 0.15) | <2.0×10<sup>-16</sup> | - | - |
| Graham *et al.* (UKB-included) | 0.28<br>(0.28, 0.28) | <2.0×10<sup>-16</sup> | 0.27<br>(0.25, 0.30) | <2.0×10<sup>-16</sup> | 0.14<br>(0.12, 0.16) | <2.0×10<sup>-16</sup> | 0.26<br>(0.22, 0.30) | <2.0×10<sup>-16</sup> |
| Willer *et al.* | 0.21<br>(0.20, 0.21) | <2.0×10<sup>-16</sup> | - | - | - | - | - | - |

**Supplementary Table 5.** Associations of lipid PGS with the corresponding circulating lipid levels stratified by verified vegetarian status in UKB participants of European and Central/South Asian ancestries (sensitivity analyses). PGS were calculated using the UKB-included GWAS from Graham *et al*., and the GWAS from Willer *et al*, respectively. The beta value indicates the effect of PGS on each blood lipid level. Abbreviations: 95% CI, 95% confident interval; PGS, polygenic scores.

| Lipids | Vegetarian |  |  | Non-vegetarian |  |  | P<sub>interaction</sub> |
| --- | --- | --- | --- | --- | --- | --- | --- |
|  | n | β (95% CI) | P-value | n | β (95% CI) | P-value |  |
| EUR |  |  |  |  |  |  |  |
| Graham *et al.* (UKB-included) |  |  |  |  |  |  |  |
| Total cholesterol | 2,275 | 0.319 (0.285, 0.354) | <2.0×10<sup>-16</sup> | 141,161 | 0.295 (0.290, 0.300) | <2.0×10<sup>-16</sup> | 0.272 |
| LDL cholesterol | 2,271 | 0.329 (0.295, 0.363) | <2.0×10<sup>-16</sup> | 140,899 | 0.306 (0.302, 0.311) | <2.0×10<sup>-16</sup> | 0.540 |
| HDL cholesterol | 2,071 | 0.300 (0.267, 0.333) | <2.0×10<sup>-16</sup> | 128,917 | 0.317 (0.312, 0.321) | <2.0×10<sup>-16</sup> | 0.334 |
| Triglycerides | 2,271 | 0.252 (0.212, 0.291) | <2.0×10<sup>-16</sup> | 141,076 | 0.280 (0.275, 0.285) | <2.0×10<sup>-16</sup> | 0.253 |
| Willer *et al.* |  |  |  |  |  |  |  |
| Total cholesterol | 2,275 | 0.267 (0.232, 0.303) | <2.0×10<sup>-16</sup> | 141,161 | 0.258 (0.253, 0.263) | <2.0×10<sup>-16</sup> | 0.840 |
| LDL cholesterol | 2,271 | 0.268 (0.233, 0.302) | <2.0×10<sup>-16</sup> | 140,899 | 0.265 (0.260, 0.270) | <2.0×10<sup>-16</sup> | 0.836 |
| HDL cholesterol | 2,071 | 0.244 (0.210, 0.278) | <2.0×10<sup>-16</sup> | 128,917 | 0.259 (0.255, 0.264) | <2.0×10<sup>-16</sup> | 0.464 |
| Triglycerides | 2,271 | 0.155 (0.116, 0.195) | 1.71×10<sup>-14</sup> | 141,076 | 0.206 (0.201, 0.210) | <2.0×10<sup>-16</sup> | 0.015 |
| CSA |  |  |  |  |  |  |  |
| Graham *et al.* (UKB-included) |  |  |  |  |  |  |  |
| Total cholesterol | 183 | 0.125 (-0.021, 0.271) | 0.095 | 1,565 | 0.171 (0.125, 0.217) | 5.69×10<sup>-13</sup> | 0.491 |
| LDL cholesterol | 183 | 0.153 (-0.009, 0.316) | 0.067 | 1,564 | 0.197 (0.151, 0.244) | <2.0×10<sup>-16</sup> | 0.160 |
| HDL cholesterol | 162 | 0.280 (0.169, 0.391) | 2.74×10<sup>-6</sup> | 1,434 | 0.217 (0.179, 0.254) | <2.0×10<sup>-16</sup> | 0.780 |
| Triglycerides | 183 | 0.264 (0.083, 0.444) | 0.005 | 1,565 | 0.264 (0.212, 0.316) | <2.0×10<sup>-16</sup> | 0.935 |

**Supplementary Table 6.**
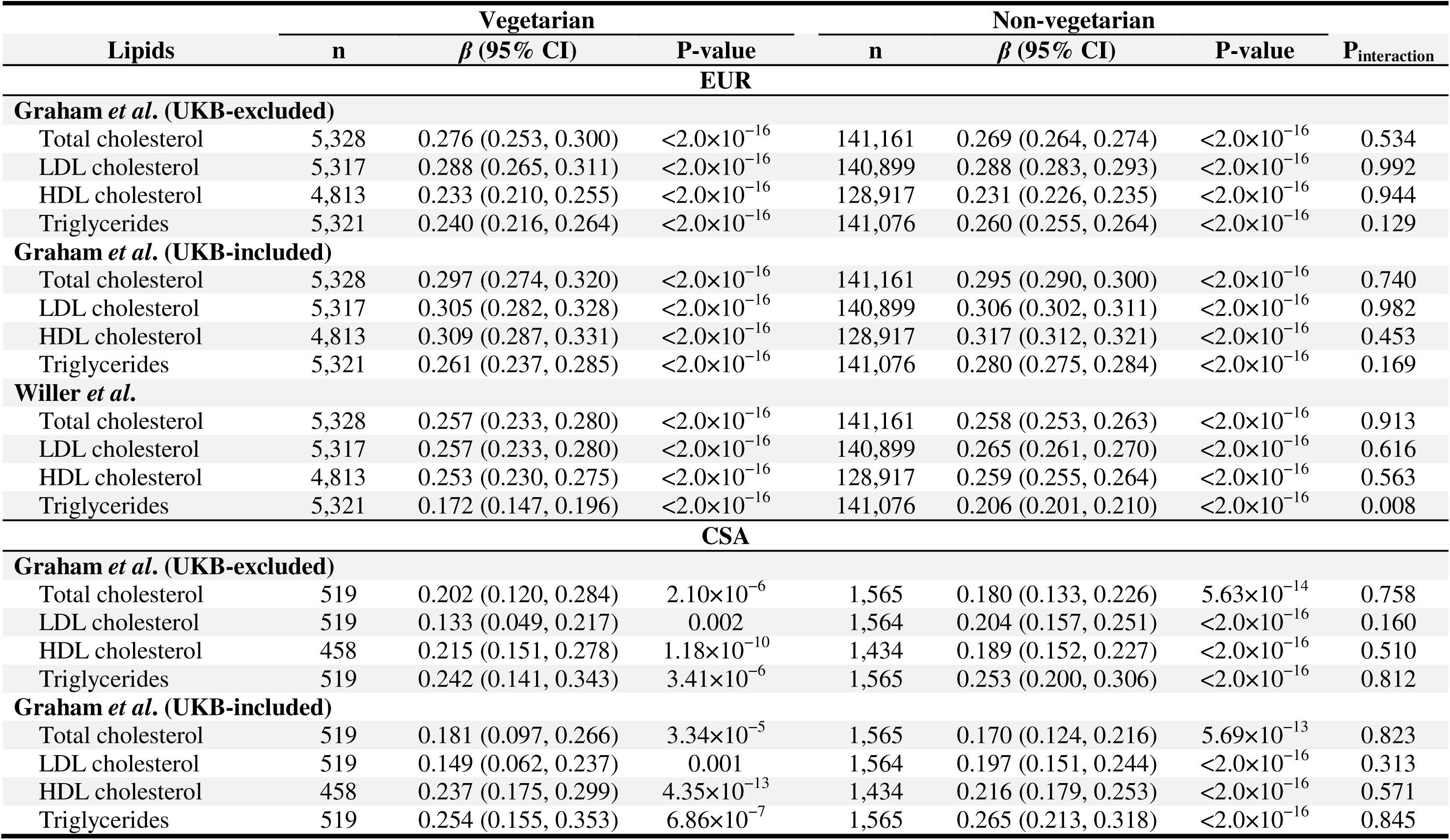
Associations of lipid PGS with the corresponding circulating lipid levels stratified by self-reported vegetarian status in UKB participants of European and Central/South Asian ancestries (sensitivity analyses). PGS were calculated using GWAS from Graham *et al*. that exclude and include UKB dataset, and from Willer *et al*, respectively. The beta value indicates the effect of PGS on each blood lipid level. The number of self-reported vegetarians and non-vegetarians was represented by n. Abbreviations: 95% CI, 95% confident interval; PGS, polygenic scores.

| Lipids | Vegetarian |  |  | Non-vegetarian |  |  | P<sub>interaction</sub> |
| --- | --- | --- | --- | --- | --- | --- | --- |
|  | n | β (95% CI) | P-value | n | β (95% CI) | P-value |  |
| EUR |  |  |  |  |  |  |  |
| Graham *et al.* (UKB-excluded) |  |  |  |  |  |  |  |
| Total cholesterol | 5,328 | 0.276 (0.253, 0.300) | <2.0×10<sup>-16</sup> | 141,161 | 0.269 (0.264, 0.274) | <2.0×10<sup>-16</sup> | 0.534 |
| LDL cholesterol | 5,317 | 0.288 (0.265, 0.311) | <2.0×10<sup>-16</sup> | 140,899 | 0.288 (0.283, 0.293) | <2.0×10<sup>-16</sup> | 0.992 |
| HDL cholesterol | 4,813 | 0.233 (0.210, 0.255) | <2.0×10<sup>-16</sup> | 128,917 | 0.231 (0.226, 0.235) | <2.0×10<sup>-16</sup> | 0.944 |
| Triglycerides | 5,321 | 0.240 (0.216, 0.264) | <2.0×10<sup>-16</sup> | 141,076 | 0.260 (0.255, 0.264) | <2.0×10<sup>-16</sup> | 0.129 |
| Graham *et al.* (UKB-included) |  |  |  |  |  |  |  |
| Total cholesterol | 5,328 | 0.297 (0.274, 0.320) | <2.0×10<sup>-16</sup> | 141,161 | 0.295 (0.290, 0.300) | <2.0×10<sup>-16</sup> | 0.740 |
| LDL cholesterol | 5,317 | 0.305 (0.282, 0.328) | <2.0×10<sup>-16</sup> | 140,899 | 0.306 (0.302, 0.311) | <2.0×10<sup>-16</sup> | 0.982 |
| HDL cholesterol | 4,813 | 0.309 (0.287, 0.331) | <2.0×10<sup>-16</sup> | 128,917 | 0.317 (0.312, 0.321) | <2.0×10<sup>-16</sup> | 0.453 |
| Triglycerides | 5,321 | 0.261 (0.237, 0.285) | <2.0×10<sup>-16</sup> | 141,076 | 0.280 (0.275, 0.284) | <2.0×10<sup>-16</sup> | 0.169 |
| Willer *et al.* |  |  |  |  |  |  |  |
| Total cholesterol | 5,328 | 0.257 (0.233, 0.280) | <2.0×10<sup>-16</sup> | 141,161 | 0.258 (0.253, 0.263) | <2.0×10<sup>-16</sup> | 0.913 |
| LDL cholesterol | 5,317 | 0.257 (0.233, 0.280) | <2.0×10<sup>-16</sup> | 140,899 | 0.265 (0.261, 0.270) | <2.0×10<sup>-16</sup> | 0.616 |
| HDL cholesterol | 4,813 | 0.253 (0.230, 0.275) | <2.0×10<sup>-16</sup> | 128,917 | 0.259 (0.255, 0.264) | <2.0×10<sup>-16</sup> | 0.563 |
| Triglycerides | 5,321 | 0.172 (0.147, 0.196) | <2.0×10<sup>-16</sup> | 141,076 | 0.206 (0.201, 0.210) | <2.0×10<sup>-16</sup> | 0.008 |
| CSA |  |  |  |  |  |  |  |
| Graham *et al.* (UKB-excluded) |  |  |  |  |  |  |  |
| Total cholesterol | 519 | 0.202 (0.120, 0.284) | 2.10×10<sup>-6</sup> | 1,565 | 0.180 (0.133, 0.226) | 5.63×10<sup>-14</sup> | 0.758 |
| LDL cholesterol | 519 | 0.133 (0.049, 0.217) | 0.002 | 1,564 | 0.204 (0.157, 0.251) | <2.0×10<sup>-16</sup> | 0.160 |
| HDL cholesterol | 458 | 0.215 (0.151, 0.278) | 1.18×10<sup>-10</sup> | 1,434 | 0.189 (0.152, 0.227) | <2.0×10<sup>-16</sup> | 0.510 |
| Triglycerides | 519 | 0.242 (0.141, 0.343) | 3.41×10<sup>-6</sup> | 1,565 | 0.253 (0.200, 0.306) | <2.0×10<sup>-16</sup> | 0.812 |
| Graham *et al.* (UKB-included) |  |  |  |  |  |  |  |
| Total cholesterol | 519 | 0.181 (0.097, 0.266) | 3.34×10<sup>-5</sup> | 1,565 | 0.170 (0.124, 0.216) | 5.69×10<sup>-13</sup> | 0.823 |
| LDL cholesterol | 519 | 0.149 (0.062, 0.237) | 0.001 | 1,564 | 0.197 (0.151, 0.244) | <2.0×10<sup>-16</sup> | 0.313 |
| HDL cholesterol | 458 | 0.237 (0.175, 0.299) | 4.35×10<sup>-13</sup> | 1,434 | 0.216 (0.179, 0.253) | <2.0×10<sup>-16</sup> | 0.571 |
| Triglycerides | 519 | 0.254 (0.155, 0.353) | 6.86×10<sup>-7</sup> | 1,565 | 0.265 (0.213, 0.318) | <2.0×10<sup>-16</sup> | 0.845 |

**Supplementary Table 7.** Associations between vegetarianism (verified) and blood lipid levels stratified by low (bottom 33%) and high (top 33%) PGS groups for the EUR and CSA cohorts. PGS were calculated using GWAS from Graham *et al*. that exclude and include UKB dataset, and from Willer *et al*, respectively. The beta value indicates the effect of verified vegetarian on each blood lipid level. Abbreviations: 95% CI, 95% confident interval; PGS, polygenic scores.

| Lipids | Low PGS |  | High PGS |  | P<sub>interaction</sub> |
| --- | --- | --- | --- | --- | --- |
|  | β (95% CI) | P-value | β (95% CI) | P-value |  |
| EUR |  |  |  |  |  |
| Total cholesterol |  |  |  |  |  |
| Graham *et al.* (UKB-excluded) | -0.146 (-0.209, -0.083) | 5.66×10<sup>-6</sup> | -0.177 (-0.249, -0.106) | 1.32×10<sup>-6</sup> | 0.605 |
| Graham *et al.* (UKB-included) | -0.172 (-0.234, -0.111) | 4.18×10<sup>-8</sup> | -0.186 (-0.258, -0.114) | 4.47×10<sup>-7</sup> | 0.366 |
| Willer *et al.* | -0.147 (-0.210, -0.084) | 4.58×10<sup>-6</sup> | -0.199 (-0.272, -0.127) | 7.93×10<sup>-8</sup> | 0.942 |
| LDL cholesterol |  |  |  |  |  |
| Graham *et al.* (UKB-excluded) | -0.169 (-0.233, -0.106) | 1.89×10<sup>-7</sup> | -0.198 (-0.270, -0.126) | 7.90×10<sup>-8</sup> | 0.384 |
| Graham *et al.* (UKB-included) | -0.169 (-0.231, -0.107) | 1.11×10<sup>-7</sup> | -0.199 (-0.272, -0.126) | 1.01×10<sup>-7</sup> | 0.411 |
| Willer *et al.* | -0.134 (-0.198, -0.070) | 4.17×10<sup>-5</sup> | -0.207 (-0.279, -0.136) | 1.48×10<sup>-8</sup> | 0.689 |
| HDL cholesterol |  |  |  |  |  |
| Graham *et al.* (UKB-excluded) | -0.164 (-0.220, -0.108) | 1.05×10<sup>-8</sup> | -0.188 (-0.253, -0.123) | 1.83×10<sup>-8</sup> | 0.95 |
| Graham *et al.* (UKB-included) | -0.138 (-0.192, -0.083) | 7.78×10<sup>-7</sup> | -0.215 (-0.279, -0.150) | 6.84×10<sup>-11</sup> | 0.326 |
| Willer *et al.* | -0.155 (-0.210, -0.099) | 6.34×10<sup>-8</sup> | -0.200 (-0.263, -0.136) | 6.66×10<sup>-10</sup> | 0.665 |
| Triglycerides |  |  |  |  |  |
| Graham *et al.* (UKB-excluded) | 0.162 (0.110, 0.213) | 6.42×10<sup>-10</sup> | 0.226 (0.147, 0.306) | 2.45×10<sup>-8</sup> | 0.491 |
| Graham *et al.* (UKB-included) | 0.156 (0.107, 0.206) | 7.15×10<sup>-10</sup> | 0.208 (0.129, 0.287) | 2.21×10<sup>-7</sup> | 0.33 |
| Willer *et al.* | 0.188 (0.135, 0.241) | 4.35×10<sup>-12</sup> | 0.179 (0.101, 0.257) | 6.17×10<sup>-6</sup> | 0.07 |
| CSA |  |  |  |  |  |
| Total cholesterol |  |  |  |  |  |
| Graham *et al.* (UKB-excluded) | 0.059 (-0.208, 0.326) | 0.665 | -0.090 (-0.384, 0.204) | 0.55 | 0.284 |
| Graham *et al.* (UKB-included) | 0.093 (-0.165, 0.351) | 0.482 | -0.032 (-0.335, 0.271) | 0.836 | 0.198 |
| LDL cholesterol |  |  |  |  |  |
| Graham *et al.* (UKB-excluded) | 0.117 (-0.157, 0.392) | 0.403 | 0.045 (-0.258, 0.349) | 0.771 | 0.352 |
| Graham *et al.* (UKB-included) | 0.167 (-0.098, 0.432) | 0.218 | 0.035 (-0.260, 0.331) | 0.814 | 0.272 |
| HDL cholesterol |  |  |  |  |  |
| Graham *et al.* (UKB-excluded) | -0.139 (-0.334, 0.055) | 0.161 | -0.186 (-0.428, 0.057) | 0.134 | 0.634 |
| Graham *et al.* (UKB-included) | -0.145 (-0.342, 0.051) | 0.148 | -0.267 (-0.510, -0.023) | 0.032 | 0.892 |
| Triglycerides |  |  |  |  |  |
| Graham *et al.* (UKB-excluded) | 0.297 (0.035, 0.559) | 0.027 | 0.178 (-0.240, 0.597) | 0.404 | 0.682 |
| Graham *et al.* (UKB-included) | 0.363 (0.088, 0.637) | 0.01 | 0.187 (-0.223, 0.597) | 0.372 | 0.58 |

**Supplementary Table 8.** Associations between vegetarianism (self-reported) and blood lipid levels stratified by low (bottom 33%) and high (top 33%) PGS groups for the EUR and CSA cohorts. PGS were calculated using GWAS from Graham *et al*. that exclude and includes the UKB dataset, and from Willer *et al*, respectively. The beta value indicates the effect of self-reported vegetarianism on each blood lipid level. Abbreviations: 95% CI, 95% confident interval; PGS, polygenic scores; EUR, European; CSA, Central/South Asian.

| Lipids | Low PGS |  | High PGS |  | P<sub>interaction</sub> |
| --- | --- | --- | --- | --- | --- |
|  | β (95% CI) | P-value | β (95% CI) | P-value |  |
| EUR |  |  |  |  |  |
| Total cholesterol |  |  |  |  |  |
| Graham *et al.* (UKB-excluded) | -0.136 (-0.178, -0.095) | 1.38×10<sup>-10</sup> | -0.164 (-0.211, -0.116) | 1.28×10<sup>-11</sup> | 0.642 |
| Graham *et al.* (UKB-included) | -0.137 (-0.178, -0.096) | 6.40×10<sup>-11</sup> | -0.176 (-0.223, -0.128) | 5.23×10<sup>-13</sup> | 0.949 |
| Willer *et al.* | -0.129 (-0.171, -0.087) | 1.88×10<sup>-9</sup> | -0.171 (-0.218, -0.123) | 2.30×10<sup>-12</sup> | 0.871 |
| LDL cholesterol |  |  |  |  |  |
| Graham *et al.* (UKB-excluded) | -0.122 (-0.164, -0.081) | 4.98E-08 | -0.168 (-0.216, -0.120) | 6.38×10<sup>-12</sup> | 0.891 |
| Graham *et al.* (UKB-included) | -0.122 (-0.164, -0.081) | 5.95×10<sup>-9</sup> | -0.170 (-0.218, -0.122) | 5.44×10<sup>-12</sup> | 0.813 |
| Willer *et al.* | -0.107 (-0.150, -0.064) | 8.80×10<sup>-7</sup> | -0.171 (-0.219, -0.123) | 2.53×10<sup>-12</sup> | 0.48 |
| HDL cholesterol |  |  |  |  |  |
| Graham *et al.* (UKB-excluded) | -0.134 (-0.171, -0.097) | 1.05×10<sup>-12</sup> | -0.152 (-0.196, -0.108) | 1.37×10<sup>-11</sup> | 0.75 |
| Graham *et al.* (UKB-included) | -0.119 (-0.155, -0.083) | 6.99×10<sup>-11</sup> | -0.168 (-0.211, -0.125) | 1.77×10<sup>-14</sup> | 0.539 |
| Willer *et al.* | -0.115 (-0.152, -0.079) | 7.23×10<sup>-10</sup> | -0.147 (-0.190, -0.104) | 2.58×10<sup>-11</sup> | 0.766 |
| Triglycerides |  |  |  |  |  |
| Graham *et al.* (UKB-excluded) | 0.102 (0.068, 0.137) | 4.14×10<sup>-9</sup> | 0.154 (0.102, 0.205) | 4.27×10<sup>-9</sup> | 0.295 |
| Graham *et al.* (UKB-included) | 0.109 (0.075, 0.142) | 1.57×10<sup>-10</sup> | 0.148 (0.096, 0.199) | 1.63×10<sup>-8</sup> | 0.143 |
| Willer *et al.* | 0.139 (0.103, 0.176) | 5.52×10<sup>-14</sup> | 0.116 (0.066, 0.166) | 5.39×10<sup>-6</sup> | 0.004 |
| CSA |  |  |  |  |  |
| Total cholesterol |  |  |  |  |  |
| Graham *et al.* (UKB-excluded) | -0.216 (-0.382, -0.050) | 0.011 | -0.060 (-0.248, 0.128) | 0.535 | 0.968 |
| Graham *et al.* (UKB-included) | -0.176 (-0.340, -0.011) | 0.037 | -0.065 (-0.258, 0.127) | 0.505 | 0.96 |
| LDL cholesterol |  |  |  |  |  |
| Graham *et al.* (UKB-excluded) | -0.061 (-0.230, 0.107) | 0.475 | -0.033 (-0.223, 0.157) | 0.731 | 0.355 |
| Graham *et al.* (UKB-included) | -0.045 (-0.210, 0.119) | 0.591 | -0.085 (-0.274, 0.104) | 0.378 | 0.358 |
| HDL cholesterol |  |  |  |  |  |
| Graham *et al.* (UKB-excluded) | -0.107 (-0.240, 0.026) | 0.115 | -0.089 (-0.242, 0.063) | 0.25 | 0.524 |
| Graham *et al.* (UKB-included) | -0.128 (-0.259, 0.002) | 0.055 | -0.132 (-0.286, 0.022) | 0.094 | 0.873 |
| Triglycerides |  |  |  |  |  |
| Graham *et al.* (UKB-excluded) | 0.175 (0.008, 0.342) | 0.04 | 0.123 (-0.128, 0.375) | 0.337 | 0.742 |
| Graham *et al.* (UKB-included) | 0.181 (0.011, 0.352) | 0.037 | 0.185 (-0.069, 0.438) | 0.154 | 0.915 |

## References

1. Cramer, H., et al., Characteristics of Americans Choosing Vegetarian and Vegan Diets for Health Reasons. J Nutr Educ Behav, 2017. 49(7): p. 561–567.e1.

2. Pilis, W., et al., Health benefits and risk associated with adopting a vegetarian diet. Rocz Panstw Zakl Hig, 2014. 65(1): p. 9–14.

3. Melina, V., W. Craig, and S. Levin, Position of the Academy of Nutrition and Dietetics: Vegetarian Diets. J Acad Nutr Diet, 2016. 116(12): p. 1970–1980.

4. Lee, K.W., et al., Effects of Vegetarian Diets on Blood Pressure Lowering: A Systematic Review with Meta-Analysis and Trial Sequential Analysis. Nutrients, 2020. 12(6).

5. Wang, F., et al., Effects of Vegetarian Diets on Blood Lipids: A Systematic Review and Meta-Analysis of Randomized Controlled Trials. J Am Heart Assoc, 2015. 4(10): p. e002408.

6. Koch, C.A., E.W. Kjeldsen, and R. Frikke-Schmidt, Vegetarian or vegan diets and blood lipids: a meta-analysis of randomized trials. Eur Heart J, 2023. 44(28): p. 2609–2622.

7. Bradbury, K.E., et al., Serum concentrations of cholesterol, apolipoprotein A-I and apolipoprotein B in a total of 1694 meat-eaters, fish-eaters, vegetarians and vegans. Eur J Clin Nutr, 2014. 68(2): p. 178–83.

8. Landry, M.J., et al., Cardiometabolic Effects of Omnivorous vs Vegan Diets in Identical Twins: A Randomized Clinical Trial. JAMA Netw Open, 2023. 6(11): p. e2344457.

9. Crowe, F.L., et al., Risk of hospitalization or death from ischemic heart disease among British vegetarians and nonvegetarians: results from the EPIC-Oxford cohort study. Am J Clin Nutr, 2013. 97(3): p. 597–603.

10. Tonstad, S., et al., Vegetarian diets and incidence of diabetes in the Adventist Health Study-2. Nutr Metab Cardiovasc Dis, 2013. 23(4): p. 292–9.

11. Vanacore, D., et al., Effect of restriction vegan diet’s on muscle mass, oxidative status, and myocytes differentiation: A pilot study. J Cell Physiol, 2018. 233(12): p. 9345–9353.

12. Ahmad, F.B. and R.N. Anderson, The Leading Causes of Death in the US for 2020. JAMA, 2021. 325(18): p. 1829–1830.

13. Pirillo, A., et al., Global epidemiology of dyslipidaemias. Nat Rev Cardiol, 2021. 18(10): p. 689–700.

14. Panagiotakos, D.B., et al., Effect of leisure time physical activity on blood lipid levels: the ATTICA study. Coron Artery Dis, 2003. 14(8): p. 533–9.

15. Patel, C.J., et al., Systematic evaluation of environmental factors: persistent pollutants and nutrients correlated with serum lipid levels. Int J Epidemiol, 2012. 41(3): p. 828–43.

16. Sun, Y., et al., Fish oil supplementation modifies the associations between genetically predicted and observed concentrations of blood lipids: a cross-sectional gene-diet interaction study in UK Biobank. Am J Clin Nutr, 2024. 120(3): p. 540–549.

17. Francis, M., et al., Gene-vegetarianism interactions in calcium, estimated glomerular filtration rate, and testosterone identified in genome-wide analysis across 30 biomarkers. PLoS Genet, 2024. 20(7): p. e1011288.

18. Bassin, S.R., J. Ferreira De Carvalho, and M. Gulati, A Review of Plant-Based Diets for Obesity Management. Endocr Pract, 2024. 30(8): p. 779–789.

19. De Biase, S.G., et al., Vegetarian diet and cholesterol and triglycerides levels. Arq Bras Cardiol, 2007. 88(1): p. 35–9.

20. Garousi, N., et al., Effects of lacto-ovo-vegetarian diet vs. standard-weight-loss diet on obese and overweight adults with non-alcoholic fatty liver disease: a randomised clinical trial. Arch Physiol Biochem, 2023. 129(4): p. 975–983.

21. Kettunen, J., et al., Genome-wide association study identifies multiple loci influencing human serum metabolite levels. Nat Genet, 2012. 44(3): p. 269–76.

22. van der Laan, S.W., et al., From lipid locus to drug target through human genomics. Cardiovasc Res, 2018. 114(9): p. 1258–1270.

23. Kathiresan, S., et al., A genome-wide association study for blood lipid phenotypes in the Framingham Heart Study. BMC Med Genet, 2007. 8 **Suppl 1**(Suppl 1): p. S17.

24. Kilpelainen, T.O., et al., Multi-ancestry study of blood lipid levels identifies four loci interacting with physical activity. Nat Commun, 2019. 10(1): p. 376.

25. de Vries, P.S., et al., Multiancestry Genome-Wide Association Study of Lipid Levels Incorporating Gene-Alcohol Interactions. Am J Epidemiol, 2019. 188(6): p. 1033–1054.

26. Bentley, A.R., et al., Multi-ancestry genome-wide gene-smoking interaction study of 387,272 individuals identifies new loci associated with serum lipids. Nat Genet, 2019. 51(4): p. 636–648.

27. Francis, M., et al., Genome-wide association study of fish oil supplementation on lipid traits in 81,246 individuals reveals new gene-diet interaction loci. PLoS Genet, 2021. 17(3): p. e1009431.

28. Ottman, R., Gene-environment interaction: definitions and study designs. Prev Med, 1996. 25(6): p. 764–70.

29. Griffin, B.A., et al., APOE4 genotype exerts greater benefit in lowering plasma cholesterol and apolipoprotein B than wild type (E3/E3), after replacement of dietary saturated fats with low glycaemic index carbohydrates. Nutrients, 2018. 10(10): p. 1524.

30. MacKay, D.S., et al., CYP7A1-rs3808607and APOEisoform associate with LDL cholesterol lowering after plant sterol consumption in a randomized clinical trial. The American journal of clinical nutrition, 2015. 102(4): p. 951–957.

31. Kim, D.S., et al., Novel gene-by-environment interactions: APOB and NPC1L1 variants affect the relationship between dietary and total plasma cholesterol. J Lipid Res, 2013. 54(5): p. 1512–20.

32. Xiang, R., et al., Recent advances in polygenic scores: translation, equitability, methods and FAIR tools. Genome Med, 2024. 16(1): p. 33.

33. Campos, A.I., et al., Boosting the power of genome-wide association studies within and across ancestries by using polygenic scores. Nature Genetics, 2023. 55(10): p. 1769–1776.

34. Domingue, B.W., et al., Interactions between polygenic scores and environments: Methodological and conceptual challenges. Sociological Science, 2020. 7: p. 465–486.

35. D’Urso, S. and L.-D. Hwang, New Insights into Polygenic Score–Lifestyle Interactions for Cardiometabolic Risk Factors from Genome-Wide Interaction Analyses. Nutrients, 2023. 15(22): p. 4815.

36. Ye, Y., et al., Interactions between enhanced polygenic risk scores and lifestyle for cardiovascular disease, diabetes, and lipid levels. Circulation: Genomic and Precision Medicine, 2021. 14(1): p. e003128.

37. Ripatti, P., et al., Polygenic Hyperlipidemias and Coronary Artery Disease Risk. Circ Genom Precis Med, 2020. 13(2): p. e002725.

38. Wu, H., et al., Polygenic Risk Score for Low-Density Lipoprotein Cholesterol Is Associated With Risk of Ischemic Heart Disease and Enriches for Individuals With Familial Hypercholesterolemia. Circ Genom Precis Med, 2021. 14(1): p. e003106.

39. Tam, C.H.T., et al., Development of genome-wide polygenic risk scores for lipid traits and clinical applications for dyslipidemia, subclinical atherosclerosis, and diabetes cardiovascular complications among East Asians. Genome Med, 2021. 13(1): p. 29.

40. Sudlow, C., et al., UK biobank: an open access resource for identifying the causes of a wide range of complex diseases of middle and old age. PLoS Med, 2015. 12(3): p. e1001779.

41. Karczewski, K.J., et al., Pan-UK Biobank GWAS improves discovery, analysis of genetic architecture, and resolution into ancestry-enriched effects. medRxiv, 2024: p. 2024.03.13.24303864.

42. Bycroft, C., et al., The UK Biobank resource with deep phenotyping and genomic data. Nature, 2018. 562(7726): p. 203–209.

43. Ritchie, S.C., et al., Quality control and removal of technical variation of NMR metabolic biomarker data in ∼120,000 UK Biobank participants. Sci Data, 2023. 10(1): p. 64.

44. Choi, S.W., T.S. Mak, and P.F. O’Reilly, Tutorial: a guide to performing polygenic risk score analyses. Nat Protoc, 2020. 15(9): p. 2759–2772.

45. Willer, C.J., et al., Discovery and refinement of loci associated with lipid levels. Nat Genet, 2013. 45(11): p. 1274–1283.

46. Graham, S.E., et al., The power of genetic diversity in genome-wide association studies of lipids. Nature, 2021. 600(7890): p. 675–679.

47. Desmond, M.A., et al., Growth, body composition, and cardiovascular and nutritional risk of 5- to 10-y-old children consuming vegetarian, vegan, or omnivore diets. Am J Clin Nutr, 2021. 113(6): p. 1565–1577.

48. Sacks, F.M., et al., Dietary Fats and Cardiovascular Disease: A Presidential Advisory From the American Heart Association. Circulation, 2017. 136(3): p. e1–e23.

49. Abumweis, S.S., R. Barake, and P.J. Jones, Plant sterols/stanols as cholesterol lowering agents: A meta-analysis of randomized controlled trials. Food Nutr Res, 2008. 52.

50. Ghavami, A., et al., Soluble Fiber Supplementation and Serum Lipid Profile: A Systematic Review and Dose-Response Meta-Analysis of Randomized Controlled Trials. Adv Nutr, 2023. 14(3): p. 465–474.

51. Yokoyama, Y., S.M. Levin, and N.D. Barnard, Association between plant-based diets and plasma lipids: a systematic review and meta-analysis. Nutr Rev, 2017. 75(9): p. 683–698.

52. Thomas, D., Methods for investigating gene-environment interactions in candidate pathway and genome-wide association studies. Annu Rev Public Health, 2010. 31: p. 21–36.

53. Tang, Y., et al., IPRS: Leveraging Gene-Environment Interaction to Reconstruct Polygenic Risk Score. Front Genet, 2022. 13: p. 801397.

54. Ni, G., et al., A Comparison of Ten Polygenic Score Methods for Psychiatric Disorders Applied Across Multiple Cohorts. Biol Psychiatry, 2021. 90(9): p. 611–620.

